# Delineating disorder-general and disorder-specific dimensions of psychopathology from functional brain networks in a developmental clinical sample

**DOI:** 10.1101/2023.03.31.23288009

**Authors:** Irene Voldsbekk, Rikka Kjelkenes, Andreas Dahl, Madelene C. Holm, Martina J. Lund, Tobias Kaufmann, Christian K. Tamnes, Ole A. Andreassen, Lars T. Westlye, Dag Alnæs

**Author notes:** Corresponding author: Full name: Irene Voldsbekk Address: NORMENT, Oslo universitetssykehus HF Klinikk psykisk helse og avhengighet Seksjon for psykoseforskning/TOP Ullevål sykehus, bygg 49 Postboks 4956 Nydalen 0424 Oslo.

## Abstract

The interplay between functional brain network maturation and psychopathology during development remains elusive. To establish the structure of psychopathology and its neurobiological mechanisms, mapping of both shared and unique functional connectivity patterns across developmental clinical populations is needed. We investigated shared associations between resting-state functional connectivity and psychopathology in children and adolescents aged 5-21 (n=1689). Specifically, we used partial least squares (PLS) to identify latent variables (LV) between connectivity and both symptom scores and diagnostic information. We also investigated associations between connectivity and each diagnosis specifically, controlling for other diagnosis categories. PLS identified five significant LVs between connectivity and symptoms, mapping onto the psychopathology hierarchy. The first LV resembled a general psychopathology factor, followed by dimensions of internalising-externalising, neurodevelopment, somatic complaints, and thought problems. Another PLS with diagnostic data revealed one significant LV, resembling a cross-diagnostic case-control pattern. The diagnosis-specific PLS identified a unique connectivity pattern for autism spectrum disorder (ASD). All LVs were associated with distinct patterns of functional connectivity. These dimensions largely replicated in an independent sample (n=420) from the same dataset, as well as to an independent cohort (n = 3504). This suggests that covariance in developmental functional brain networks supports transdiagnostic dimensions of psychopathology.

## 1 Introduction

Establishing the structure of psychopathology and its underlying neurobiological mechanisms is a critical step towards personalised approaches in mental health research and care. The high rate of comorbidity between diagnoses challenges the utility of traditional case-control designs and motivates novel strategies for clinical phenotyping such as transdiagnostic assessment of psychopathology dimensions (Caspi et al., 2014; Insel et al., 2010). Existing diagnostic categories do not map onto disease-specific neurobiological substrates (Insel & Cuthbert, 2015), and many of the detected abnormalities in both genetics (Hindley et al., 2022; Lahey et al., 2011; Pettersson et al., 2016; Roelfs et al., 2021), brain structure (Goodkind et al., 2015; Opel et al., 2020a) and brain function (Elliott et al., 2018; McTeague et al., 2017; McTeague et al., 2020; Sha et al., 2019) are shared across disorders. Investigation into the neurobiological substrates of distinct symptom dimensions may therefore elucidate the brain-based underpinnings of mental disorders.

Childhood and adolescence are characterised by large scale reorganisation and maturation of the brain and its functional networks (Paus et al., 2008; Power et al., 2010). Given that mental disorders often first manifest during this time (Caspi et al., 2020; Kessler et al., 2007), aberrant functional network development may represent a key aetiological component in mental disorders (Casey et al., 2014; Paus et al., 2008). Indeed, while sensory and motor regions and their associated functional networks typically are fully developed by late childhood, the association cortex, and implicated functional networks such as the default mode network (DMN), take longer to mature. This might leave these brain regions vulnerable to emerging psychopathology during neurodevelopment (Sydnor et al., 2021). To identify biologically informed dimensions of psychopathology, investigating associations between functional brain networks and psychopathology during childhood and adolescence is imperative.

Psychopathology is increasingly conceptualised as a hierarchical structure (Caspi et al., 2014; Kotov et al., 2017; Lahey et al., 2017). This hierarchy consists of a general psychopathology factor as the highest order, reflecting a general vulnerability to psychopathology, followed by increasingly narrow dimensions, such as internalising and externalising. These reflect anxious and depressive symptoms, and aggressive, rule-breaking, and hyperactive symptoms, respectively. A neurodevelopmental factor is also often included to reflect autistic-like traits and symptoms of attention deficit hyperactivity disorder (ADHD). For example, recent work in the Adolescent Brain Cognitive Development (ABCD) cohort (Casey et al., 2018) derived five dimensions of psychopathology (i.e. internalising, externalising, neurodevelopmental, detachment, and somatoform) using exploratory factor analysis on symptom data (Michelini et al., 2019). Similar psychopathology dimensions have been derived from both symptom data (Karcher et al., 2021) and diagnostic data (Lees et al., 2021) and then associated with patterns of functional connectivity obtained from resting-state functional magnetic resonance imaging (rs-fMRI). However, these studies derived dimensions of psychopathology from symptom data or diagnostic information in isolation, and only afterwards associated them with functional connectivity. To identify brain-based dimensions of psychopathology, the functional brain networks should inform the estimation of psychopathology dimensions per se.

Doing exactly this, studies have identified symptom dimensions by finding their maximal correlation with functional connectivity in youth aged 8-22 (Xia et al., 2018) and preadolescents aged 9-11 (i.e. the ABCD cohort) (Kebets et al., 2023). In youth, dimensions of mood, psychosis, fear, and externalisation symptoms exhibited both unique and a shared pattern of connectivity. In ABCD, dimensions derived from structural and functional brain patterns simultaneously resembled a general psychopathology factor along with internalising-externalising, neurodevelopmental, somatoform, and detachment dimensions. Although this work is promising with respect to identifying biologically informed dimensions of psychopathology, the investigation of mental health symptoms in population-based studies may not generalise to clinical populations (Vanes & Dolan, 2021). While investigation in young, representative cohorts is essential to understand putative developmental mechanisms relevant for psychopathological vulnerability, it is equally important to map the relevance of these findings to individuals already diagnosed with a mental disorder. Mapping of disorder-general and disorder-specific patterns in clinical populations is needed to elucidate the underlying neurobiological mechanisms of psychopathology.

Patterns of connectivity related to symptoms of anxiety, irritability, and ADHD were replicated across two independent clinical samples of children and adolescents (Linke et al., 2021). Specifically, this study identified one dimension consisting of all three domains, while the second dimension captured shared aspects of irritability and ADHD, and the third was specific to anxiety. This indicates clinically relevant disorder-general (i.e. shared across disorders) and disorder-specific effects in functional networks of children and adolescents. Moreover, it points to the possibility of decomposing irritability, a symptom shared between anxiety and ADHD, into disorder-specific and common components based on patterns of brain connectivity. However, this study did not investigate connectivity patterns related to broad transdiagnostic symptom dimensions but maintained a focus limited to anxiety, irritability, and attention problems. Moreover, the degree of overlap between functional networks linked to symptom dimensions and those related to diagnosis remain to be determined.

In the current study, we aimed to investigate dimensions linking functional connectivity and psychopathology in a sample of children and adolescents where the majority had at least one diagnosed mental disorder. We used partial least squares (PLS) (Krishnan et al., 2011), a multivariate technique that identifies shared associations across two high-dimensional matrices. This enables identification of dimensions of psychopathology derived from connectivity patterns in functional brain networks. Specifically, we wanted to highlight similarities and differences across dimensions derived from symptom data vs diagnostic classifications in the same sample. To do this, we investigated associations between functional connectivity and a) symptom scores, and b) diagnostic information. In addition, we investigated associations between functional connectivity and c) each diagnosis specifically, controlling for other diagnosis categories. To ensure robustness, we ran our analysis in a discovery subsample and then validated these findings in a replication sample from the same cohort. Finally, we validated the symptom dimensions of our findings in the ABCD cohort by formally comparing our results to the work by Kebets and colleagues (Kebets et al., 2023).

## 2 Material and methods

### 2.1 Sample

The sample was recruited from New York City, USA to participate in the Healthy Brain Network (HBN) (Alexander et al., 2017), a cohort consisting of children and adolescents aged 5-21. Participants were recruited through “community-referred recruitment,” meaning advertisements to encourage participation of families who have concerns about in the mental health of their child. Exclusion criteria were: any present acute safety concerns (e.g., being a danger to oneself or to others), cognitive or behavioural impairments hindering participation (e.g., being nonverbal) or medical concerns that likely will confound brain-related findings. The participants underwent a comprehensive assessment of biological and socio-environmental factors, in addition to diagnostic evaluation by qualified health personnel. After quality control and data cleaning (see below), the final sample for our analyses consisted of 1880 participants (721 females). This sample was then split into a discovery sample (80%, n=1689) and a replication sample (20%, n=420), matching the two subsamples on scanner location, age, sex, and diagnosis categories. See Figure S1 for a sample flow chart. Sample demographics are provided in Figure 1.

**Figure 1.**
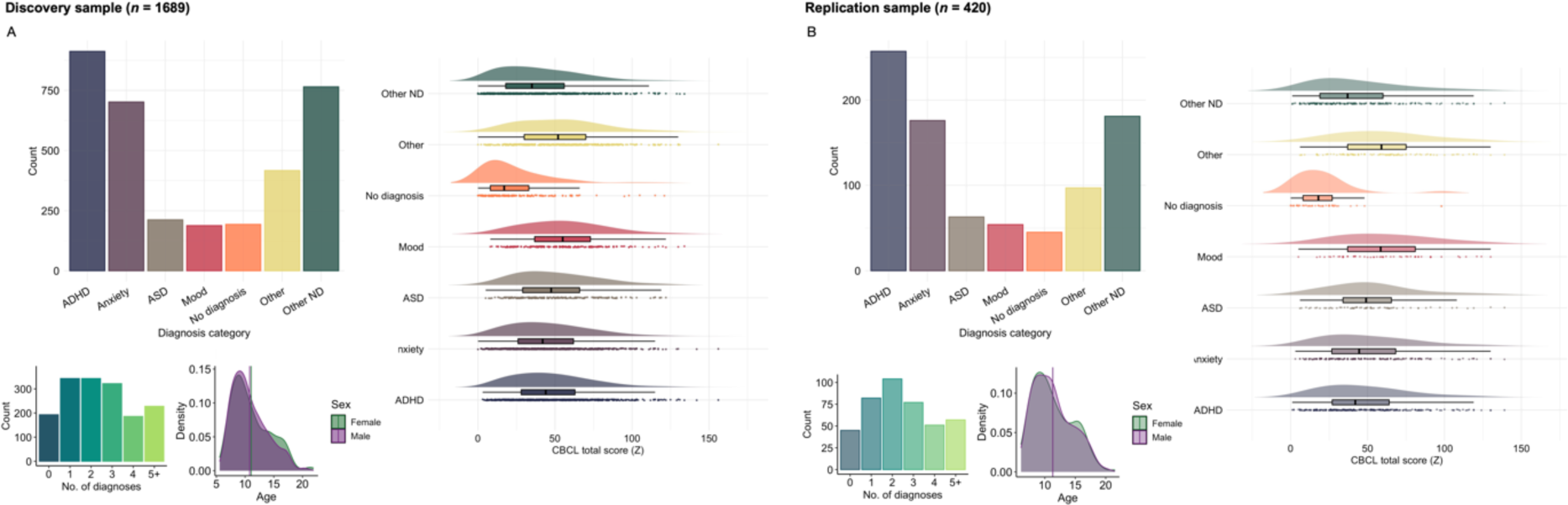
Sample distributions of diagnosis categories (more than one per individual possible), clinical symptoms, comorbidity, age, and sex. The lines indicate mean age for each sex. **A.** Discovery sample (n=1689). **B.** Replication sample (n=420). CBCL; child behaviour checklist. ADHD; attention-deficit hyperactivity disorder. ASD; autism spectrum disorder. ND; neurodevelopmental.

### 2.2 Clinical measures

Symptom scores were obtained from the Child Behaviour Checklist (CBCL) (Achenbach & Rescorla, 2001), which assesses emotional, behavioural, and social problems in children by parent report. Parents scored their children on 113 items as either 0 (“Not true”), 1 (“Somewhat or sometimes true”) or 2 (“Very true or often true”). The responses to these items result in eight syndrome measures, previously found to be the best-fitting model for data obtained from both general and clinical populations (Achenbach & Rescorla, 2001; Ivanova et al., 2007): Anxious/Depressed, Withdrawn/Depressed, Somatic Complaints, Social Problems, Thought Problems, Attention Problems, Rule-Breaking Behaviour, and Aggressive Behaviour.

Diagnostic information was obtained by a computerised version of the Schedule for Affective Disorders and Schizophrenia – Childreńs version (KSADS) (Kaufman et al., 1997), which is a clinician-administered semi-structured psychiatric interview based on DSM-5. Based on this interview and review of all other collected materials, a consensus regarding clinical diagnosis was made by a team of licensed clinicians. We then categorised diagnoses as either “ADHD”, “ASD”, “anxiety disorders”, “mood disorders”, “other neurodevelopmental disorders”, “other disorders” or “no diagnosis”. Most participants had more than one diagnosis.

Of those with complete MRI data (see below), 1992 participants had available both diagnostic data and symptom data. Participants with more than 10% missing symptom data were excluded (n=112). For the remaining participants (n=1880), missing values were imputed with knnimpute in MATLAB (MathWorks, 2020).

### 2.3 MRI pre-processing

We accessed rs-fMRI and T_1_-weighted structural MRI for the current study. MRI data were acquired at four different sites: a mobile scanner at Staten Island (SI), Rutgers University Brain Imaging Centre, Citigroup Biomedical Imaging Centre (CBIC) and Harlem CUNY Advanced Science Research Centre. A detailed overview of the MRI protocol is available elsewhere (http://fcon_1000.projects.nitrc.org/indi/cmi_healthy_brain_network/MRI%20Protocol.html). T1-weighted MRI data (n=3334) were processed using FreeSurfer v. 7.1.0 (Fischl, 2012) and quality controlled using the MRIQC classifier (Esteban et al., 2017). For participants with more than one T_1_-weighted scan, we selected the sequence with the best estimated quality, as previously described (Voldsbekk et al., 2023).

For individuals with sufficient structural MRI image quality (n=3213), we submitted rs-fMRI images for pre-processing along the following pipeline. We applied FSL MCFLIRT (Jenkinson et al., 2002) for motion correction, high-pass temporal filtering (cut-off: 100), spatial smoothing (FWHM: 6) and distortion correction as part of FEAT (Woolrich et al., 2001). The rs-fMRI images were also registered to the structural image using FLIRT (Jenkinson et al., 2002) and boundary-based registration (Greve & Fischl, 2009). Next, for additional removal of artefacts and noise, we performed non-aggressive ICA-AROMA (Pruim, Mennes, Buitelaar, et al., 2015; Pruim, Mennes, van Rooij, et al., 2015) and FIX (Griffanti et al., 2014; Salimi-Khorshidi et al., 2014) with a threshold of 20. During this procedure, 595 participants were excluded due to missing data or insufficient image quality. As an additional step, quality control of the raw rs-fMRI images was performed using MRIQC. Estimations of temporal signal-to-noise ratio (tSNR) and mean framewise displacement (FD), as calculated by MRIQC, were used as covariates in subsequent analyses.

### 2.4 Network analysis

To increase reproducibility, nodes were estimated from the Schaefer parcellation with 100 parcels and 7 networks (Schaefer et al., 2018). These networks include visual A, visual B, visual C, auditory, somatomotor A, somatomotor B, language, salience A, salience B, control A, control B, control C, default A, default B, default C, dorsal attention A and dorsal attention B. As an additional quality check of the estimated parcels, participants with data in less than 60% of voxels for each parcel were excluded (n=290). An overview of percentage missing data in each parcel is shown in Figure S2. To check that 60% is a reasonable threshold, balancing exclusion of participants vs completeness of voxel data, time series correlations were computed in the subset of participants with no missing data between the full parcel time series (i.e., from 100% of voxels) and parcel time series based on 60% of the voxels of that parcel (removing those voxels most frequently missing). The correlation between the full 100% parcel and the 60% parcel time series was high for every parcel (all higher than .87, see Figure S3). Parcel timeseries were then imported to FSLNets (https://fsl.fmrib.ox.ac.uk/fsl/fslwiki/FSLNets), as implemented in MATLAB (MathWorks, 2020), for estimation of edges (n=2328). In this step, we calculated the partial correlations between nodes using L2-norm ridge regression, as these are considered a better measure of direct connectivity strength (Marrelec et al., 2006). Finally, edges were z-transformed using Fisher’s transformation and we extracted the upper triangle of the correlation matrix for further analysis, yielding 4,950 unique edges reflecting the connection strength between nodes for each participant.

### 2.5 Partial-least squares

To assess shared associations between edges (i.e., functional connectivity strength between two brain regions) and mental health data in the discovery sample, we used PLS Application (Krishnan et al., 2011), as this toolbox affords a straightforward implementation of contrasts. This approach yields latent variables (LV) reflecting maximal covariance across both matrices. See figure 2 for an overview of the PLS analysis pipeline. Briefly, the PLS analysis estimates a cross-covariance matrix between imaging and behavioural data. This matrix is then inputted to singular value decomposition, yielding a total number of LVs corresponding to the number of behavioural variables. For each of these LVs, we get a singular value and the weights of each imaging and behavioural variable, as well as for each subject. The significance of LVs was assessed using permutation testing (n=5000). Then, the stability of edges for each significant LV was estimated using bootstrapping with replacement (n=1000), thresholding at |pseudo-z|>3| (McIntosh & Lobaugh, 2004) for significance. Loadings onto each LV was then extracted as the correlation between weights on each LV and the original data. First, to investigate symptom-based patterns, we ran a rotated behavioural PLS with z-transformed symptom data as behavioural variables. Next, entering diagnostic information (one column for each diagnosis: 1 as having the diagnosis, 0 as not – more than one possible per participant) as behavioural variables, we used the same rotated behavioural PLS approach to decompose data into putative specific and shared disorder dimensions (i.e., diagnosis-based patterns). Then, to test for diagnosis-specific patterns explicitly, we ran non-rotated behavioural PLS, in which associations between edges and each diagnostic category was tested. This test was run for each diagnosis category separately, while controlling for all other diagnosis categories (see Table S1 for an overview of contrasts).

**Figure 2.**
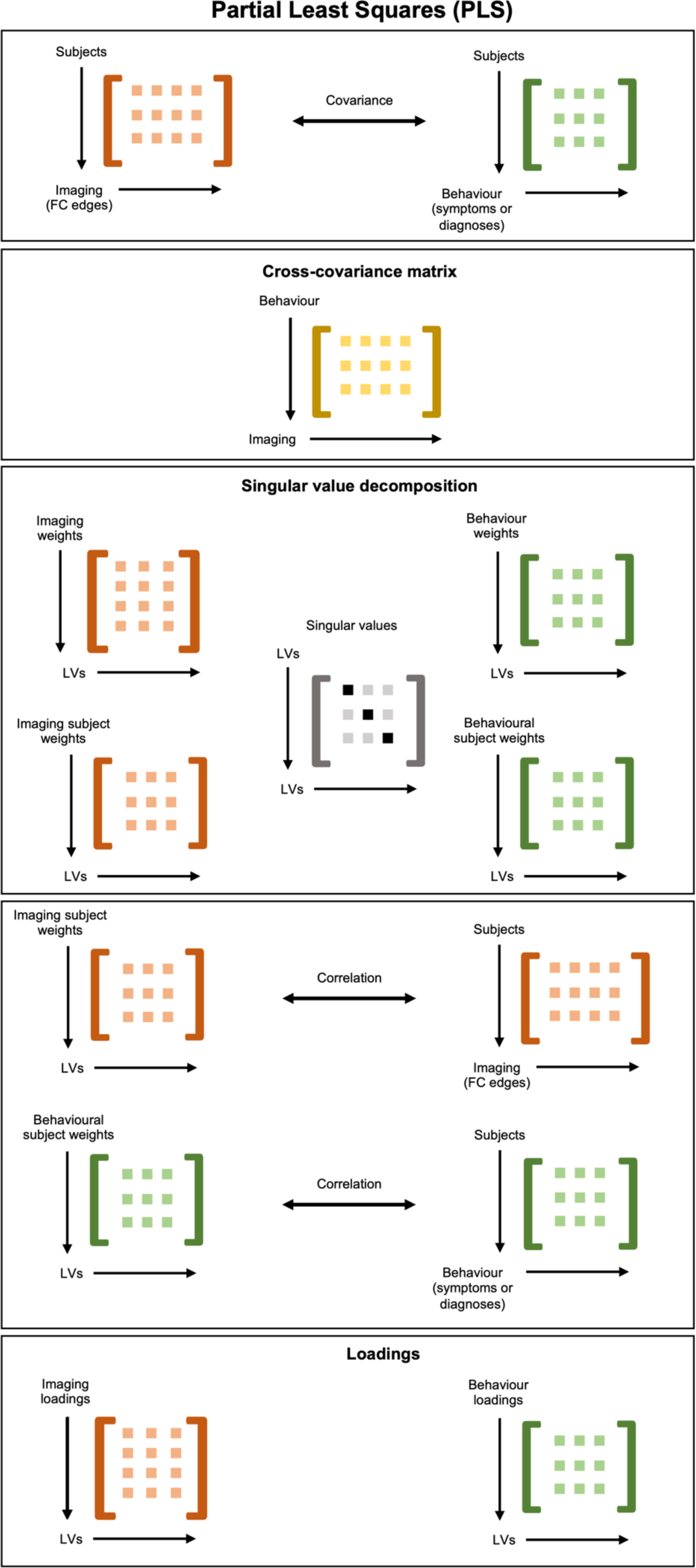
Illustration of the PLS analysis pipeline. The cross-covariance matrix between imaging data and behavioural data is estimated. This matrix is then inputted to singular value decomposition, yielding singular values for each LV, as well as imaging, behavioural, and subject weights. Then, loadings onto each LV are calculated as the correlation between subject weights and the original imaging and behavioural data, respectively. PLS; partial least squares. LV; latent variable.

Prior to running PLS, edges were adjusted for sex, age, tSNR, FD, and scanner site. PLS was run using Spearmańs rank correlation. Significant LVs were then plotted using R version 4.1.2 (https://cran.r-project.org). To aid in the visualisation and interpretation of the high dimensional connectivity patterns, edges were summarised across networks for significant LVs. To investigate the loading of nodes, we also estimated the nodal loading strength across the connectivity matrix for each connectivity pattern identified by PLS using the Brain Connectivity Toolbox (Rubinov & Sporns, 2010) in MATLAB. To obtain a more detailed overview of each connectivity pattern, we plotted nodal strength and edge strength using BrainNet Viewer (Xia et al., 2013).

### 2.6 Consistency across age, sex, ethnicity, socio-economic status, intelligence, and medication use

Given that aberrant brain development may represent a key aetiological component in mental disorders (Casey et al., 2014; Paus et al., 2008), we assessed whether the shared associations between edges and symptom data in the discovery sample differed as a function of age or sex. To do this, we reran the symptom-based PLS without adjusting edges for age and sex. These results revealed similar patterns of covariation for all LVs except LV4, which was not found (Figure S4). However, all LVs exhibited highly correlated feature weights across overlapping dimensions (Table S2). To examine whether the shared associations identified were generalisable across ethnic groups, we plotted the correlations between edges and symptoms by ethnic group (Figure S5). Similarly, to examine whether the shared associations identified were generalisable across levels of socioeconomic status (SES), we plotted the correlations by median-split of household income, as a proxy for SES (Figure S6). To examine whether the shared associations identified were generalisable across levels of intelligence, we plotted the correlations by full scale IQ split into ± 70 (Figure S7). To examine whether the shared associations identified were generalisable across current use of psychiatric medication or not, we plotted the correlations by current use vs no use (yes/no; 310 participants reported yes) (Figure S8). Finally, symptom loadings and connectivity loadings from the symptom-based PLS were regressed against each diagnosis category separately, with “no diagnosis” as a reference group. We also regressed loadings against number of diagnoses, interpreting the latter as a proxy of cross-diagnostic vulnerability. All associations were adjusted for age, age^2^, and sex.

### 2.7 Validation in replication sample

To test whether the results were robust and reliable, we repeated the PLS analysis in the replication sample. Akin to previous work (Linke et al., 2021), weights estimated in each subsample were then multiplied with input data from the other subsample to derive subject weights for participants whose data was not part of the model estimations. Replication was determined as the Pearson’s correlation of the derived subject weights for each dataset with those estimated in the corresponding subset. To establish significance of the correlations, we ran permutations (n=5000) and results were considered replicable if correlations in both directions were significant.

### 2.8 Validation in independent cohort

To test whether the results also generalise to other cohorts, we formally tested the replicability of previous work in the ABCD cohort (Kebets et al., 2023) to the current sample. Specifically, we repeated the replication procedure described above, this time applying weights from the ABCD dataset to input data from HBN and correlated this product with subject weights from our symptom-based PLS analysis. Of note, as this work utilised the Schaefer parcellation with 400 parcels and submitted these to PCA prior to running PLS, we first decomposed the HBN data by multiplying 400 parcellated HBN data with PCA weights estimated in the ABCD analysis.

## 3 Results

### 3.1 Symptom-based dimensions

Based on the scree plot of percent cross-block covariance explained (Figure S9), we selected the first six LVs in the symptom-based PLS in the discovery sample for further analysis. Of these, five were significant (r=.72, p=.045; r=.65, p=.026; r=.75, p=.009; r=.71, p=.031; r=.62, p=.003, respectively; Figure S10). Each LV represents a distinct pattern that relates a weighted set of symptoms to a weighted set of functional brain network connections. Inspection of the most heavily weighted symptoms for each LV revealed that they resemble the psychopathology hierarchy: the first LV resembled a general psychopathology factor (see Figure 3A), while the remaining four represented increasingly narrow dimensions (see Figure 4). Specifically, LV2 was related to internalising-externalising, LV3 to neurodevelopment, LV4 to somatic complaints, and LV6 to thought problems.

**Figure 3.**
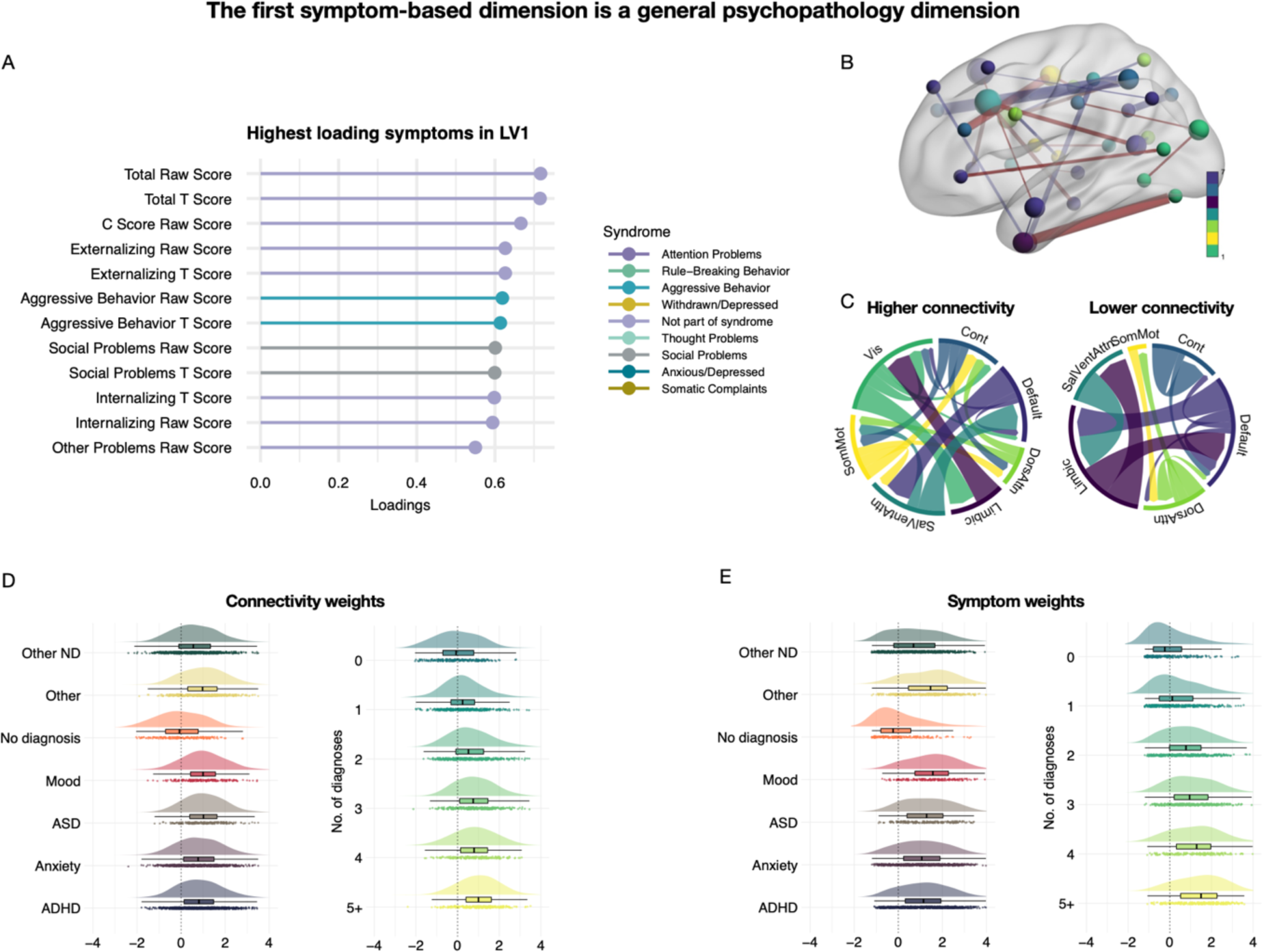
The first dimension of shared associations between functional connectivity and clinical symptoms resembled a general psychopathology factor. **A.** The highest loading symptoms of this dimension. Loadings reflect correlations between LV weights and original data. **B.** Strength of edges and nodes that contributed to this dimension. Edges are coloured red for higher connectivity and blue for lower connectivity. Nodes are coloured based on network membership. **C.** Both increased and reduced connectivity in specific edges contributed. Magnitude in this plot reflects summarised edge strength across each network. **E-F.** Connectivity and symptom loadings across diagnostic categories (left) and number of comorbidities (right). In these plots, the data is centred around the mean of no diagnosis. LV; latent variable. ADHD; attention-deficit hyperactivity disorder. ASD; autism spectrum disorder. ND; neurodevelopmental. Vis; visual network. SomMot; somatomotor network. SalVentAttn; salience network. Cont; control network. Default; default mode network. DorsAttn; dorsal attention network.

**Figure 4.**
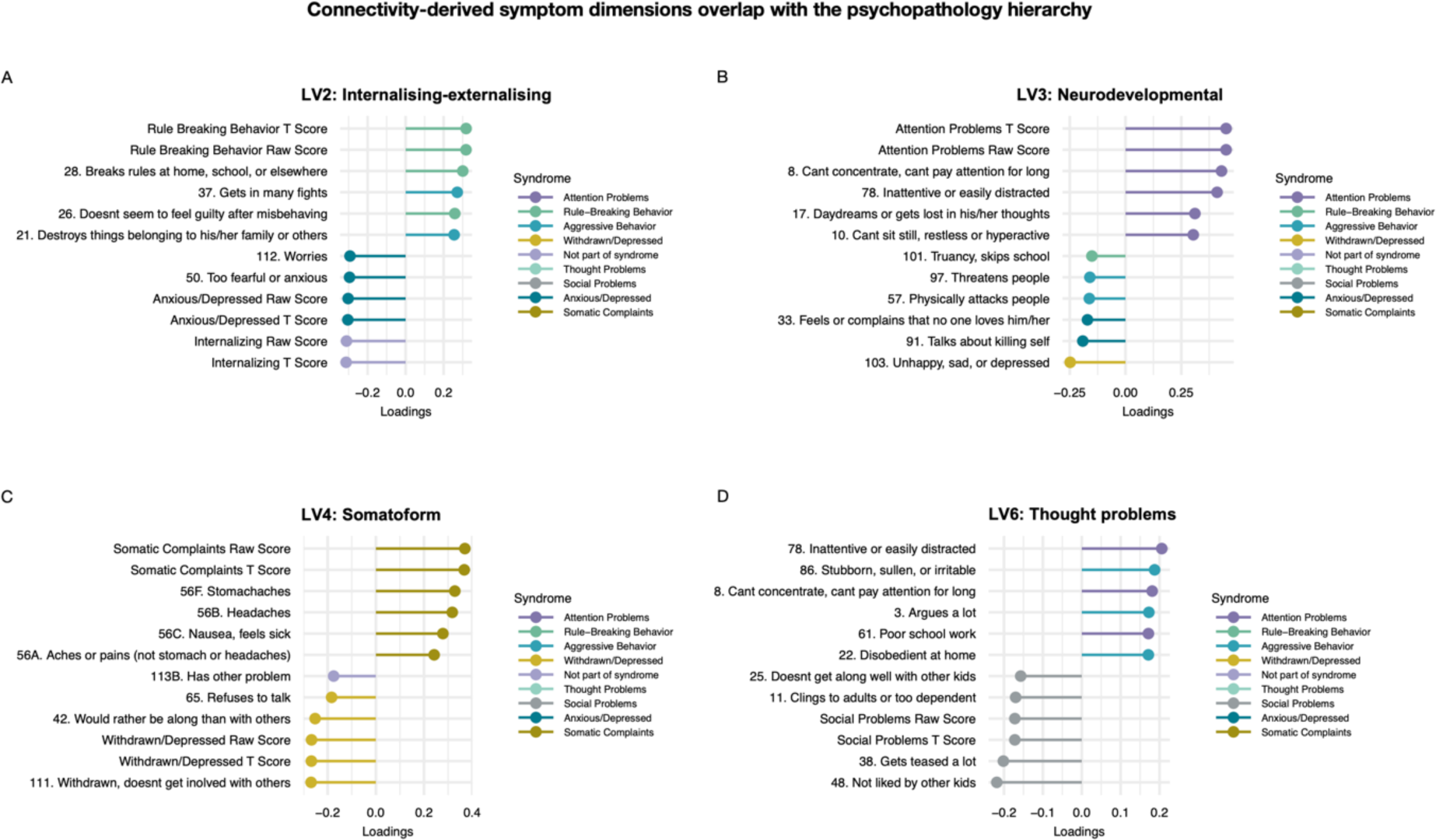
Dimensions of shared associations between functional connectivity and clinical symptoms map onto the hierarchical structure of psychopathology. **A.** LV2 map onto symptoms of higher externalisation and lower internalisation. **B.** LV3 map onto symptoms of neurodevelopmental problems. **C.** LV4 map onto symptoms of higher somatic complaints and lower withdrawn/depressive symptoms. **D.** LV6 map onto symptoms of higher thought problems. LV; latent variable.

These psychopathology dimensions were identified by their shared associations with specific patterns of connectivity. For each dimension, these patterns were widely distributed across functional networks (see Figure 3 and S11). LV1 was related to weaker connectivity between the salience and limbic network, and between the limbic network and DMN, as well as within the control network (Figure 3B-C). In addition, LV1 was related to stronger connectivity between the limbic and visual network and between the salience network and DMN. The nodes with strongest loading on this pattern implicated the control network, DMN, and salience network. LV2 was related to stronger within- and between-network connectivity in the visual network and lower connectivity within and between the salience network and DMN. The strongest nodes in LV2 were in the DMN, control and DA network (Figure S11). LV3 was related to stronger connectivity between the somatomotor and visual network and to lower connectivity between the limbic network and control network. The strongest nodes were distributed across the DMN, control, and visual network. LV4 was related to weaker connectivity between the DA and control network, stronger connectivity between the visual and limbic network, and within the DA and somatomotor network. The strongest nodes were in the DA network. LV6 was related to weaker connectivity between the limbic and visual network and between the visual network and the DA network, as well as between the somatomotor network and DMN. The strongest nodes were in the control network and DA network.

### 3.2 Diagnosis-based dimensions

The rotated diagnosis-based PLS identified one significant LV (r=.68, p=.009). As shown in Figure 5A, this pattern resembled a general case vs control pattern across all diagnoses. Partly consistent with the general psychopathology pattern of the symptom-based LV1, a higher loading on this LV (i.e. having a diagnosis) entailed stronger connectivity between the limbic and visual and salience network, in addition to weaker connectivity between the salience network and DMN (Figure 5B-C). The strongest nodes were distributed across all these networks.

**Figure 5.**
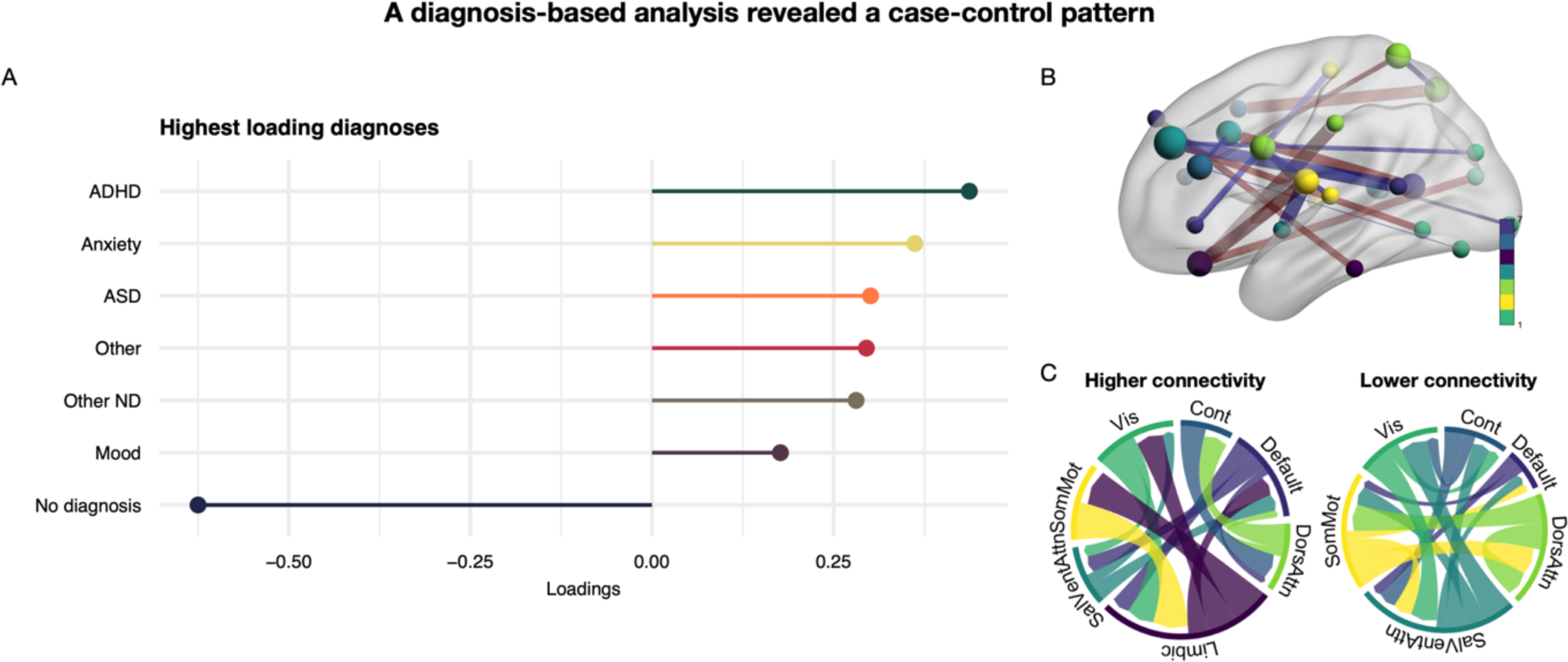
One dimension of shared associations between functional connectivity and diagnosis categories, resembling a cross-diagnostic case-control difference. **A.** The diagnosis dimension reflected a pattern across all diagnostic categories vs no diagnosis. **B.** Strength of edges and nodes that contributed to this dimension. Edges are coloured red for higher connectivity and blue for lower connectivity. Nodes are coloured based on network membership. **C.** Both increased and reduced connectivity in specific edges contributed. Magnitude in this plot reflects summarised edge strength across each network. ADHD; attention-deficit hyperactivity disorder. ND; neurodevelopmental. ASD; autism spectrum disorder. Vis; visual network. SomMot; somatomotor network. SalVentAttn; salience network. Cont; control network. Default; default mode network. DorsAttn; dorsal attention network.

### 3.3 Diagnosis-specific patterns

The non-rotated diagnosis-specific PLS, which tested each diagnosis category separately while controlling for all other diagnosis categories, identified a unique connectivity pattern for ASD (r=.44, p=.012). As shown in Figure 6, the ASD-specific pattern was widely distributed, including weaker connectivity within the somatomotor network and between the DA network and visual network. The strongest nodes implicated the salience, visual, and DA network. In addition, the non-rotated diagnosis-specific PLS identified a unique pattern of no diagnosis vs all diagnoses (r=.56, p=.012) (Figure S12). This pattern implicated increased connectivity between the somatomotor, and salience and DA network, in addition to lower connectivity between the visual and limbic network. The strongest nodes implicated in this pattern were in the salience and somatoform network. The remaining diagnosis categories did not show disorder-specific patterns of functional connectivity.

**Figure 6.**
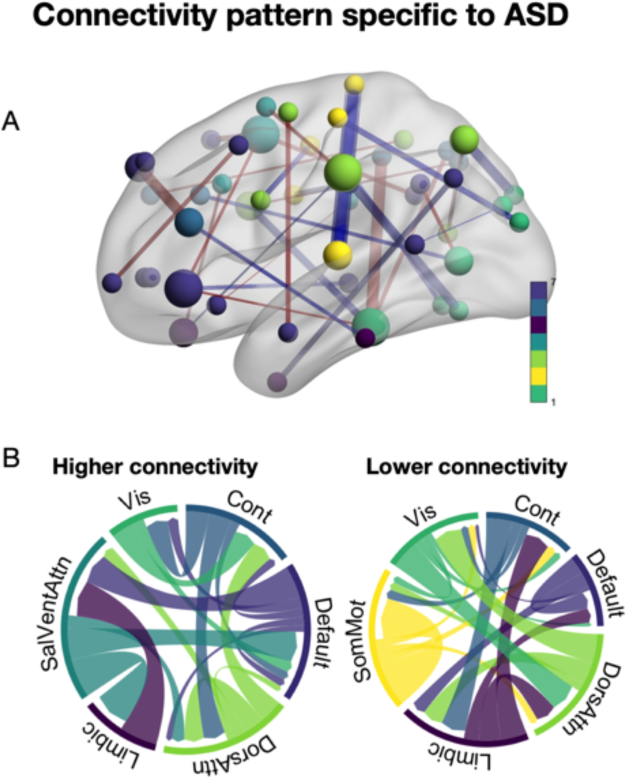
ASD was the only diagnosis category exhibiting a unique pattern of connectivity **A.** Strength of edges and nodes specific to ASD. Edges are coloured red for higher connectivity and blue for lower connectivity. Nodes are coloured based on network membership. **B.** Both increased and reduced connectivity in specific edges contributed. Magnitude in this plot reflects summarised edge strength across each network. ASD; autism spectrum disorder. Vis; visual network. SomMot; somatomotor network. SalVentAttn; salience network. Cont; control network. Default; default mode network. DorsAttn; dorsal attention network.

### 3.4 Symptom-based pattern evident across diagnostic boundaries

To understand the distribution of symptom dimensions in more detail, we plotted them against diagnosis categories (Figure S13). As shown in Figure 3E-F, there was a consistency between a higher degree of comorbidity and higher loading on LV1. We also observed expected variation in symptom loading with respect to specific diagnoses, such as patients with ADHD and ASD loading more highly on the neurodevelopmental dimension (LV3; Figure S13). On the internalising-externalising dimension (LV2), mood disorder, ASD, and anxiety disorder loaded more negatively, consistent with increasing symptoms of internalising being typical for these diagnosis categories. In addition, linear models revealed that all diagnosis categories showed higher symptom and connectivity loadings on LV1 compared to having no diagnosis (Table S3). There was also a significant linear association with the number of diagnoses for both symptom and connectivity loadings on LV1 (Table S4 and S5). This was true when including “no diagnosis” in the model or not, suggesting that this effect was not driven by case-control effects.

### 3.5 Validation in replication sample

The item weights from the symptom-based PLS were validated in the replication sample (Figure S14). For LV1-LV3, the correlations between replication and discovery and vice versa were high (r=.91-.99, all p=.001), while the correlations for LV4 and LV6 were lower (r=.13-.20, all p=.001, and r=.10-.15, p<.016, respectively). The connectivity weights for LV2, LV3 and LV6 were also significantly correlated (r=.08-.18, p<.025), while LV1 and LV4 did not replicate (r=.04-.14, p=.001-.18) (Figure S14). Of note, the association between connectivity weights for LV1 and LV4 were correlated between replication-derived discovery weights and original discovery weights, while the opposite direction was not.

For the diagnosis-based PLS and the diagnosis-specific PLS, a similar pattern emerged. The diagnosis weights in the diagnosis-based PLS were replicated across the discovery and replication samples (both r=.99, p=.001), while the connectivity weights were not (r>.06, p=.002-.094) (Figure S15). For the ASD-specific pattern derived in the diagnosis-specific PLS, the connectivity weights were validated across samples (r=.20-.21, both p=.001), while no diagnosis-specific connectivity weights were not replicated (both r=.04, p=.07-.20) (Figure S16).

### 3.6 Validation in independent cohort

The application of ABCD-derived PLS weights to HBN data revealed replication of all five symptom LVs in the symptom-based PLS across cohorts. As shown in Figure 7, the LV1-LV3 symptom weights were highly correlated (r=.86-.98, all p=.001), while LV4 and LV6 exhibited lower correlations (r=.12, p=.001, and r=.21, p=.001, respectively). The connectivity weights did not replicate (r=.001-.02,p>.18).

**Figure 7.**
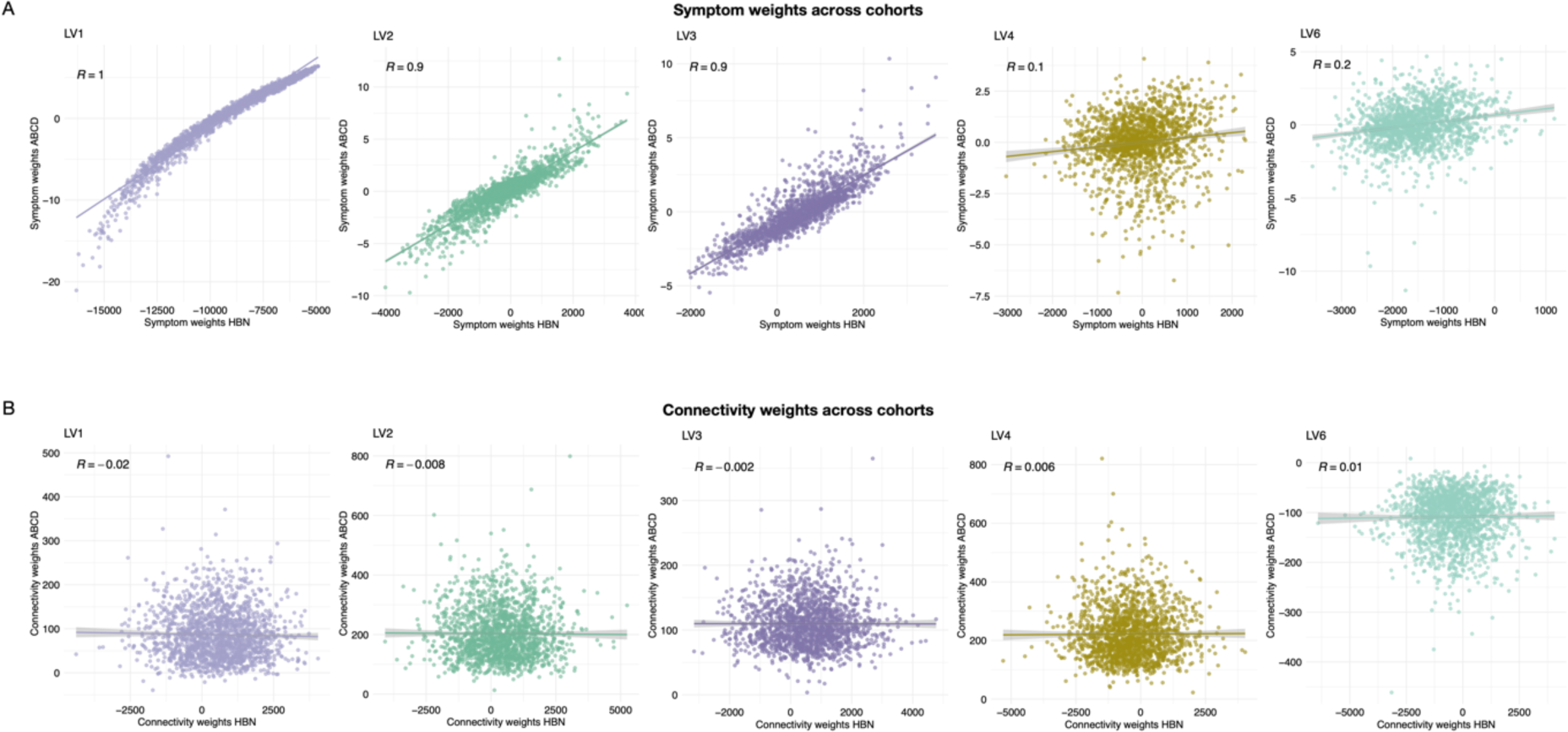
Correlations between PLS weights derived in the ABCD cohort and in the current sample (i.e., HBN). **A.** Significant associations across ABCD and HBN for symptom weights. **B.** No significant associations for connectivity weights. PLS; partial least squares. ABCD; adolescent brain cognitive development cohort. HBN; healthy brain network cohort.

## 4 Discussion

Through shared associations between mental health data and functional connectivity, the current study delineated shared and unique patterns in child and adolescent functional brain networks. We found that dimensions of clinical symptoms map onto specific patterns of brain connectivity, aligned with the psychopathology hierarchy consisting of a general psychopathology factor and increasingly narrow dimensions. The rotated decomposition of diagnostic data (i.e. the diagnosis-based PLS) revealed one significant dimension, implicating a cross-diagnostic pattern only. The disorder-specific tests revealed specific patterns of connectivity related to ASD and no diagnosis (i.e., a disorder-general effect), but not for any other diagnosis. For the symptom-based dimensions, we found that increased comorbidity was consistently related to both increased symptom burden and increased connectivity aberrations. Critically, these clinical patterns were largely replicable in an independent sample from the same dataset, as well as in an independent cohort, supporting the robustness and generalisability of our findings. Consistent with previous work (Linke et al., 2021), the connectivity patterns were not replicable to the same extent. Taken together, these results indicate that compared to diagnostic classifications in isolation, transdiagnostic and symptom-based dimensions of psychopathology are more closely mapped to the functional networks of the brain during the formative years of childhood and adolescence.

The clinical dimensions revealed by shared associations between functional connectivity and symptoms in the current study adhere to the hierarchical structure of psychopathology, implicating a general psychopathology factor, followed by dimensions of internalising-externalising, neurodevelopment, somatic complaints, and thought problems. PLS derives orthogonal LVs, leaving dimensions independent. Capturing internalisation-externalisation as the second latent pattern is consistent with previous work (Kebets et al., 2023; Linke et al., 2021). Indeed, we did not only detect overlapping symptom-based dimensions as those previously identified by Kebets and colleagues (Kebets et al., 2023), we were able to replicate them in our sample. This replication across samples is striking, suggesting generalisable patterns of functional connectivity-psychopathology associations, and strongly supporting the conceptualisation of general population vs clinical population as existing on a continuum. Interestingly, given the overlap between the current functional connectivity-derived symptom dimensions and CBCL subscale syndromes derived from symptom data alone, it appears that the psychopathology hierarchy is represented in functional networks during development.

In contrast to our findings, Linke and colleagues (Linke et al., 2021) identified a dimension specific to anxiety symptoms, which did not emerge in the current study. Neither did we detect a pattern specific to having an anxiety diagnosis. This discrepancy may reflect diversity in the range of symptoms and diagnostic groups included. Indeed, in the current study, the diagnostic range was broader than in the study by Linke and colleagues (Linke et al., 2021), while the symptom domains assessed by Xia and colleagues (Xia et al., 2018) were more closely mapped to adult psychopathology than to child symptomology, which may explain their differences. Although the age range is largely overlapping between the current sample and the sample used by Xia and colleagues, the current sample had a mean age of 10.5 years, while the previous study had a mean age of 15.82 years. Consistent with this difference in age, the diagnoses and symptoms prevalent in the current sample were neurodevelopmental and early emerging psychopathologies, such as attention problems, autism, and anxiety, while the sample used by Xia et al (2018) present symptomatology more closely resembling distributions seen in adolescent and adult samples, including all those childhood categories, but also markedly higher prevalence of symptoms of mood disorders and emerging psychosis (Alnaes et al., 2018). It is not unreasonable to expect this difference in symptom distribution to yield differences in the clinical dimensions derived in this work compared to the previous study. In addition, the current study included CBCL summary scores, alongside item scores. This may have influenced the way our model structured the associations, which may also explain why we obtain different results.

Alterations in functional connectivity of the DMN have previously been implicated in several neurological and mental disorders (van den Heuvel & Sporns, 2019). In addition, DMN connectivity has been linked to general psychopathology (Elliott et al., 2018; Karcher et al., 2021; Kebets et al., 2023; Sato et al., 2018). In line with this, DMN nodes were some of the strongest loading nodes and edges of the general psychopathology factor (i.e. symptom-based LV1) in the current study. This factor was further characterised by a distributed pattern involving weaker connectivity between the limbic network and salience network and DMN. It also implicated stronger connectivity between DMN and salience network, and between the control network and somatomotor network. In line with this, the limbic, salience, fronto-parietal and sensorimotor networks are also implicated in general psychopathology (Vanes & Dolan, 2021).

The connectivity pattern related to a general psychopathology factor in the ABCD sample (Kebets et al., 2023) implicated increased connectivity between the DMN and salience network, which was also a key finding in the current sample. However, this pattern was not replicable across cohorts when formally comparing them. The lack of generalisability of connectivity patterns across cohorts is consistent with previous replication attempts across cohorts (Linke et al., 2021). This study showed that while connectivity-informed clinical dimensions were replicable across two cohorts, the connectivity patterns themselves were less so (Linke et al., 2021). The authors attributed this to a “many-to-one” mapping between neural and clinical variables, which may also explain the lack of overlap in specific connectivity patterns identified in the current work compared to previous work.

The diagnosis-based analysis revealed only one significant dimension, resembling a cross-diagnostic case-control pattern. This pattern was characterised by no diagnosis exhibiting the highest loading, with all the other diagnosis categories exhibiting smaller associations in the opposite direction. If, instead, connectivity patterns specific to each diagnosis were detectable, we would have expected this test to reveal several dimensions (i.e., LVs), each consisting of loadings from one (or a few) diagnoses. In addition, we identified a no diagnosis vs all diagnoses specific pattern, representing an inverse cross-diagnostic case-control pattern. Although the weighting of each diagnosis category differed between these two analyses, the overarching connectivity patterns for these dimensions revealed inverse overlap. Indeed, although the symptom-based general psychopathology dimension exhibited a more distributed connectivity pattern, there was also some overlap between this pattern and cross-diagnostic case-control pattern. Together, these patterns implicate a distributed connectivity pattern implicating several key networks, such as the salience network and the limbic network, in separating no diagnosis from having a diagnosis.

ASD was the only diagnosis category exhibiting a detectable unique pattern of connectivity. This pattern was widely distributed, implicating altered connectivity within several brain networks. Hyperconnectivity within several large-scale brain networks has previously been implicated in ASD (Uddin et al., 2019). The distributed nature of the ASD-specific pattern identified in the current study, including both increased and reduced connectivity within and between several networks, is in line with the notion that both hyperconnectivity and hypoconnectivity may underlie ASD (Kana et al., 2011). Importantly, the finding that ASD was the only diagnosis group exhibiting a unique connectivity pattern has implications for our understanding of the neurobiological substrates of ASD, but also for our understanding of psychopathology and ASD in this landscape more broadly. The current work supports the understanding that rather than belonging in the general psychopathology domain, ASD likely represent a separate neurodevelopmental dimension (Opel et al., 2020b; Ronald, 2019).

Although our findings provide several new insights into the link between functional brain connectivity and the structure of childhood psychopathology, some limitations should be noted. First, functional connectivity results are known to be influenced by methodological choices (Li et al., 2021; Sala-Llonch et al., 2019; Shirer et al., 2015), complicating the identification of robust and replicable results. To increase replicability of the current work, we relied on an established parcellation scheme (Schaefer et al., 2018). Critically, we also validated our findings in both an independent sample from the same cohort, as well as in an independent cohort. Second, several functional connectivity patterns identified implicated the limbic network, a network known to be sensitive to susceptibility artefacts and reduced signal (Khatamian et al., 2016). Although we did additional measures to reduce the influence of reduced signal on our analysis, we cannot completely rule out that our results are influenced by this confound. Third, as the sample consisted of mainly children, and most of them with at least one mental disorder, motion was an issue. To ameliorate this influence, we used the MRIQC classifier to exclude participants with insufficient image quality, cleaned data using FIX and AROMA, and regressed out measures of image quality and motion from the data.

Fourth, the cross-sectional nature of the study design prohibits any conclusion to be drawn with respect to the within-person temporal dynamics of any identified pattern. Fifth, given that the symptom data was continuous, and the diagnostic analysis separated the sample into dichotomous groups, the power to extract maximal covariance between symptoms and functional connectivity was better than that for the diagnostic analysis. As such, these results cannot be directly compared. Finally, the sample was enriched with children diagnosed with ADHD and other neurodevelopmental conditions. Although this may be representative of a developmental clinical sample, it may not generalise to other clinical populations. For example, this has implications for the comparison of the current results to other studies investigating a derived general psychopathology factor. However, given that we could replicate previous work in the current sample, our findings seem to generalise to other populations, which is a strength of the current study.

## Conclusions

The current work found that dimensions of psychopathology derived from clinical symptoms were associated with specific patterns of functional connectivity in the developing brain, while ASD was the only diagnostic category to exhibit such a specific pattern. This contributes to a growing body of evidence in favour of dimensional and transdiagnostic classifications of psychopathology (Vanes & Dolan, 2021). In this classification, neurodevelopmental conditions such as ASD may possess specific abnormalities in functional connectivity networks above and beyond those related to general psychopathology. This has implications for the pursuit of individualised brain-based surrogate markers in mental health research and care, which in turn may lead to improved prevention and intervention of mental disorders.

## Data Availability

The data that forms the basis of this work were obtained from the open access Healthy Brain Network (http://fcon_1000.projects.nitrc.org/indi/cmi_healthy_brain_network/).
The code used in the study will be made available upon publication in a public repository (Open Science Framework) (https://osf.io/gt9dk/).

https://osf.io/gt9dk/

http://fcon_1000.projects.nitrc.org/indi/cmi_healthy_brain_network/

## Acknowledgements

This project was funded by research grants from the Research Council of Norway (Grant Nos. L.T.W: 249795, T.K: 276082, 323961. C.K.T: 288083, 323951), the South-Eastern Norway Regional Health Authority (Grant Nos. L.T.W: 2014097, 2015073, 2016083, 2018076, 2019101. C.K.T: 2019069, 2021070, 2023012, 500189. D.A: 2019107, 2020086. O.A.A: 223273), the Norwegian ExtraFoundation for Health and Rehabilitation (L.T.W: Grant No. 2015/FO5146), KG Jebsen Stiftelsen, ERA-Net Cofund through the ERA PerMed project IMPLEMENT, and the European Research Council under the European Union s Horizon 2020 research and Innovation program (L.T.W: ERC StG Grant No. 802998. O.A.A: H2020 RIA grant # 847776).

The work was performed on the Service for Sensitive Data (TSD) platform, owned by the University of Oslo, operated, and developed by the TSD service group at the University of Oslo IT-Department (USIT). Computations were also performed using resources provided by UNINETT Sigma2—the National Infrastructure for High Performance Computing and Data Storage in Norway.

## Notes

### Competing Interest Statement

O.A.A. is a consultant to cortechs.ai and has received speakers honorarium from Janssen, Lundbeck, Sunovion.

### Author Declarations

The data that forms the basis of this work were obtained from the open access Healthy Brain Network (http://fcon_1000.projects.nitrc.org/indi/cmi_healthy_brain_network/).

### Summary of Updates

The new version of the manuscript includes out-of-sample validation.

## References

Achenbach, T. M., & Rescorla, L. A. (2001). Manual for the ASEBA school-age forms & profiles: an integrated system of multi-informant assessment Burlington, VT: University of Vermont. Research Center for Children, Youth, & Families, 1617.

Alexander, L. M., Escalera, J., Ai, L., Andreotti, C., Febre, K., Mangone, A., Vega-Potler, N., Langer, N., Alexander, A., & Kovacs, M. (2017). An open resource for transdiagnostic research in pediatric mental health and learning disorders. Scientific data, 4(1), 1–26.

Alnaes, D., Kaufmann, T., Doan, N. T., Cordova-Palomera, A., Wang, Y., Bettella, F., Moberget, T., Andreassen, O. A., & Westlye, L. T. (2018). Association of Heritable Cognitive Ability and Psychopathology With White Matter Properties in Children and Adolescents. JAMA Psychiatry, 75(3), 287–295. https://doi.org/10.1001/jamapsychiatry.2017.4277

Casey, B. J., Cannonier, T., Conley, M. I., Cohen, A. O., Barch, D. M., Heitzeg, M. M., Soules, M. E., Teslovich, T., Dellarco, D. V., & Garavan, H. (2018). The adolescent brain cognitive development (ABCD) study: imaging acquisition across 21 sites. Developmental Cognitive Neuroscience, 32, 43–54.

Casey, B. J., Oliveri, M. E., & Insel, T. (2014). A neurodevelopmental perspective on the research domain criteria (RDoC) framework. Biol Psychiatry, 76(5), 350–353. https://doi.org/10.1016/j.biopsych.2014.01.006

Caspi, A., Houts, R. M., Ambler, A., Danese, A., Elliott, M. L., Hariri, A., Harrington, H., Hogan, S., Poulton, R., Ramrakha, S., Rasmussen, L. J. H., Reuben, A., Richmond-Rakerd, L., Sugden, K., Wertz, J., Williams, B. S., & Moffitt, T. E. (2020). Longitudinal Assessment of Mental Health Disorders and Comorbidities Across 4 Decades Among Participants in the Dunedin Birth Cohort Study. JAMA network open, 3(4), e203221-e203221. https://doi.org/10.1001/jamanetworkopen.2020.3221

Caspi, A., Houts, R. M., Belsky, D. W., Goldman-Mellor, S. J., Harrington, H., Israel, S., Meier, M. H., Ramrakha, S., Shalev, I., Poulton, R., & Moffitt, T. E. (2014). The p Factor: One General Psychopathology Factor in the Structure of Psychiatric Disorders? Clin Psychol Sci, 2(2), 119–137. https://doi.org/10.1177/2167702613497473

Elliott, M. L., Romer, A., Knodt, A. R., & Hariri, A. R. (2018). A Connectome-wide Functional Signature of Transdiagnostic Risk for Mental Illness. Biological Psychiatry, 84(6), 452–459. https://doi.org/10.1016/j.biopsych.2018.03.012

Esteban, O., Birman, D., Schaer, M., Koyejo, O. O., Poldrack, R. A., & Gorgolewski, K. J. (2017). MRIQC: Advancing the automatic prediction of image quality in MRI from unseen sites. PLoS ONE, 12(9), e0184661. https://doi.org/10.1371/journal.pone.0184661

Fischl, B. (2012). FreeSurfer. NeuroImage, 62(2), 774–781. https://doi.org/10.1016/j.neuroimage.2012.01.021

Goodkind, M., Eickhoff, S. B., Oathes, D. J., Jiang, Y., Chang, A., Jones-Hagata, L. B., Ortega, B. N., Zaiko, Y. V., Roach, E. L., Korgaonkar, M. S., Grieve, S. M., Galatzer-Levy, I., Fox, P. T., & Etkin, A. (2015). Identification of a Common Neurobiological Substrate for Mental Illness. JAMA Psychiatry, 72(4), 305–315. https://doi.org/10.1001/jamapsychiatry.2014.2206

Greve, D. N., & Fischl, B. (2009). Accurate and robust brain image alignment using boundary-based registration. NeuroImage, 48(1), 63–72. https://doi.org/10.1016/j.neuroimage.2009.06.060

Griffanti, L., Salimi-Khorshidi, G., Beckmann, C. F., Auerbach, E. J., Douaud, G., Sexton, C. E., Zsoldos, E., Ebmeier, K. P., Filippini, N., Mackay, C. E., Moeller, S., Xu, J., Yacoub, E., Baselli, G., Ugurbil, K., Miller, K. L., & Smith, S. M. (2014). ICA-based artefact removal and accelerated fMRI acquisition for improved resting state network imaging. NeuroImage, 95, 232–247. https://doi.org/10.1016/j.neuroimage.2014.03.034

Hindley, G., Frei, O., Shadrin, A. A., Cheng, W., O’Connell, K. S., Icick, R., Parker, N., Bahrami, S., Karadag, N., Roelfs, D., Holen, B., Lin, A., Fan, C. C., Djurovic, S., Dale, A. M., Smeland, O. B., & Andreassen, O. A. (2022). Charting the Landscape of Genetic Overlap Between Mental Disorders and Related Traits Beyond Genetic Correlation. American Journal of Psychiatry, 179(11), 833–843. https://doi.org/10.1176/appi.ajp.21101051

[Record #1022 is using a reference type undefined in this output style.]

Insel, T. R., & Cuthbert, B. N. (2015). Brain disorders? precisely. Science, 348(6234), 499–500.

Ivanova, M. Y., Achenbach, T. M., Dumenci, L., Rescorla, L. A., Almqvist, F., Weintraub, S., Bilenberg, N., Bird, H., Chen, W. J., Dobrean, A., Döpfner, M., Erol, N., Fombonne, E., Fonseca, A. C., Frigerio, A., Grietens, H., Hannesdóttir, H., Kanbayashi, Y., Lambert, M., Larsson, B., Leung, P., Liu, X., Minaei, A., Mulatu, M. S., Novik, T. S., Oh, K. J., Roussos, A., Sawyer, M., Simsek, Z., Steinhausen, H.-C., Metzke, C. W., Wolanczyk, T., Yang, H.-J., Zilber, N., Zukauskiene, R., & Verhulst, F. C. (2007). Testing the 8-Syndrome Structure of the Child Behavior Checklist in 30 Societies. Journal of Clinical Child & Adolescent Psychology, 36(3), 405–417. https://doi.org/10.1080/15374410701444363

Jenkinson, M., Bannister, P., Brady, M., & Smith, S. (2002). Improved optimization for the robust and accurate linear registration and motion correction of brain images. NeuroImage, 17(2), 825–841. https://doi.org/10.1016/S1053-8119(02)91132-8

Kana, R. K., Libero, L. E., & Moore, M. S. (2011). Disrupted cortical connectivity theory as an explanatory model for autism spectrum disorders. Physics of Life Reviews, 8(4), 410–437. https://doi.org/10.1016/j.plrev.2011.10.001

Karcher, N. R., Michelini, G., Kotov, R., & Barch, D. M. (2021). Associations Between Resting-State Functional Connectivity and a Hierarchical Dimensional Structure of Psychopathology in Middle Childhood. Biological Psychiatry: Cognitive Neuroscience and Neuroimaging, 6(5), 508–517. https://doi.org/10.1016/j.bpsc.2020.09.008

Kaufman, J., Birmaher, B., Brent, D., Rao, U. M. A., Flynn, C., Moreci, P., Williamson, D., & Ryan, N. (1997). Schedule for Affective Disorders and Schizophrenia for School-Age Children-Present and Lifetime Version (K-SADS-PL): Initial Reliability and Validity Data. Journal of the American Academy of Child & Adolescent Psychiatry, 36(7), 980–988. https://doi.org/10.1097/00004583-199707000-00021

Kebets, V., Piguet, C., Chen, J., Ooi, L. Q. R., Kirschner, M., Siffredi, V., Misic, B., Yeo, B. T., & Bernhardt, B. (2023). Multimodal neural correlates of childhood psychopathology. bioRxiv, 2023.2003.2002.530821.

Kessler, R. C., Angermeyer, M., Anthony, J. C., De Graaf, R., Demyttenaere, K., Gasquet, I., De Girolamo, G., Gluzman, S., Gureje, O., & Haro, J. M. (2007). Lifetime prevalence and age-of-onset distributions of mental disorders in the World Health Organization’s World Mental Health Survey Initiative. World psychiatry, 6(3), 168.

Khatamian, Y. B., Golestani, A. M., Ragot, D. M., & Chen, J. J. (2016). Spin-Echo Resting-State Functional Connectivity in High-Susceptibility Regions: Accuracy, Reliability, and the Impact of Physiological Noise. Brain Connectivity, 6(4), 283–297. https://doi.org/10.1089/brain.2015.0365

Kotov, R., Krueger, R. F., Watson, D., Achenbach, T. M., Althoff, R. R., Bagby, R. M., Brown, T. A., Carpenter, W. T., Caspi, A., & Clark, L. A. (2017). The Hierarchical Taxonomy of Psychopathology (HiTOP): A dimensional alternative to traditional nosologies. Journal of abnormal psychology, 126(4), 454.

Krishnan, A., Williams, L. J., McIntosh, A. R., & Abdi, H. (2011). Partial Least Squares (PLS) methods for neuroimaging: A tutorial and review. NeuroImage, 56(2), 455–475. https://doi.org/10.1016/j.neuroimage.2010.07.034

Lahey, B. B., Krueger, R. F., Rathouz, P. J., Waldman, I. D., & Zald, D. H. (2017). A hierarchical causal taxonomy of psychopathology across the life span. Psychological Bulletin, 143(2), 142–186. https://doi.org/10.1037/bul0000069

Lahey, B. B., Van Hulle, C. A., Singh, A. L., Waldman, I. D., & Rathouz, P. J. (2011). Higher-Order Genetic and Environmental Structure of Prevalent Forms of Child and Adolescent Psychopathology. Archives of general psychiatry, 68(2), 181–189. https://doi.org/10.1001/archgenpsychiatry.2010.192

Lees, B., Squeglia, L. M., McTeague, L. M., Forbes, M. K., Krueger, R. F., Sunderland, M., Baillie, A. J., Koch, F., Teesson, M., & Mewton, L. (2021). Altered Neurocognitive Functional Connectivity and Activation Patterns Underlie Psychopathology in Preadolescence. Biological Psychiatry: Cognitive Neuroscience and Neuroimaging, 6(4), 387–398. https://doi.org/10.1016/j.bpsc.2020.09.007

Li, X., Ai, L., Giavasis, S., Jin, H., Feczko, E., Xu, T., Clucas, J., Franco, A., Sólon Heinsfeld, A., & Adebimpe, A. (2021). Moving beyond processing and analysis-related variation in neuroscience. bioRxiv, 2021.2012.2001.470790.

Linke, J. O., Abend, R., Kircanski, K., Clayton, M., Stavish, C., Benson, B. E., Brotman, M. A., Renaud, O., Smith, S. M., Nichols, T. E., Leibenluft, E., Winkler, A. M., & Pine, D. S. (2021). Shared and Anxiety-Specific Pediatric Psychopathology Dimensions Manifest Distributed Neural Correlates. Biological Psychiatry, 89(6), 579–587. https://doi.org/10.1016/j.biopsych.2020.10.018

Marrelec, G., Krainik, A., Duffau, H., Pélégrini-Issac, M., Lehéricy, S., Doyon, J., & Benali, H. (2006). Partial correlation for functional brain interactivity investigation in functional MRI. NeuroImage, 32(1), 228–237.

MathWorks. (2020). MATLAB. In

McIntosh, A. R., & Lobaugh, N. J. (2004). Partial least squares analysis of neuroimaging data: applications and advances. NeuroImage, 23, S250–S263.

McTeague, L. M., Huemer, J., Carreon, D. M., Jiang, Y., Eickhoff, S. B., & Etkin, A. (2017). Identification of Common Neural Circuit Disruptions in Cognitive Control Across Psychiatric Disorders. American Journal of Psychiatry, 174(7), 676–685. https://doi.org/10.1176/appi.ajp.2017.16040400

McTeague, L. M., Rosenberg, B. M., Lopez, J. W., Carreon, D. M., Huemer, J., Jiang, Y., Chick, C. F., Eickhoff, S. B., & Etkin, A. (2020). Identification of common neural circuit disruptions in emotional processing across psychiatric disorders. American Journal of Psychiatry, 177(5), 411–421.

Michelini, G., Barch, D. M., Tian, Y., Watson, D., Klein, D. N., & Kotov, R. (2019). Delineating and validating higher-order dimensions of psychopathology in the Adolescent Brain Cognitive Development (ABCD) study. Translational Psychiatry, 9(1), 261. https://doi.org/10.1038/s41398-019-0593-4

Opel, N., Goltermann, J., Hermesdorf, M., Berger, K., Baune, B. T., & Dannlowski, U. (2020a). Cross-Disorder Analysis of Brain Structural Abnormalities in Six Major Psychiatric Disorders: A Secondary Analysis of Mega- and Meta-analytical Findings From the ENIGMA Consortium. Biological Psychiatry, 88(9), 678–686. https://doi.org/10.1016/j.biopsych.2020.04.027

Opel, N., Goltermann, J., Hermesdorf, M., Berger, K., Baune, B. T., & Dannlowski, U. (2020b). Cross-disorder analysis of brain structural abnormalities in six major psychiatric disorders: a secondary analysis of mega-and meta-analytical findings from the ENIGMA consortium. Biological Psychiatry, 88(9), 678–686.

Paus, T., Keshavan, M., & Giedd, J. N. (2008). Why do many psychiatric disorders emerge during adolescence? Nat Rev Neurosci, 9(12), 947–957. https://doi.org/10.1038/nrn2513

Pettersson, E., Larsson, H., & Lichtenstein, P. (2016). Common psychiatric disorders share the same genetic origin: a multivariate sibling study of the Swedish population. Molecular Psychiatry, 21(5), 717–721. https://doi.org/10.1038/mp.2015.116

Power, J. D., Fair, D. A., Schlaggar, B. L., & Petersen, S. E. (2010). The development of human functional brain networks. Neuron, 67(5), 735-748.

Pruim, R. H. R., Mennes, M., Buitelaar, J. K., & Beckmann, C. F. (2015). Evaluation of ICA-AROMA and alternative strategies for motion artifact removal in resting state fMRI. NeuroImage, 112, 278–287. https://doi.org/10.1016/j.neuroimage.2015.02.063

Pruim, R. H. R., Mennes, M., van Rooij, D., Llera, A., Buitelaar, J. K., & Beckmann, C. F. (2015). ICA-AROMA: A robust ICA-based strategy for removing motion artifacts from fMRI data. NeuroImage, 112, 267–277. https://doi.org/10.1016/j.neuroimage.2015.02.064

Roelfs, D., Alnæs, D., Frei, O., van der Meer, D., Smeland, O. B., Andreassen, O. A., Westlye, L. T., & Kaufmann, T. (2021). Phenotypically independent profiles relevant to mental health are genetically correlated. Transl Psychiatry, 11(1), 202. https://doi.org/10.1038/s41398-021-01313-x

Ronald, A. (2019). Editorial: The psychopathology p factor: will it revolutionise the science and practice of child and adolescent psychiatry? [https://doi.org/10.1111/jcpp.13063]. Journal of Child Psychology and Psychiatry, 60(5), 497-499. https://doi.org/10.1111/jcpp.13063

Rubinov, M., & Sporns, O. (2010). Complex network measures of brain connectivity: uses and interpretations. NeuroImage, 52(3), 1059–1069.

Sala-Llonch, R., Smith, S. M., Woolrich, M., & Duff, E. P. (2019). Spatial parcellations, spectral filtering, and connectivity measures in fMRI: Optimizing for discrimination. Human Brain Mapping, 40(2), 407–419.

Salimi-Khorshidi, G., Douaud, G., Beckmann, C. F., Glasser, M. F., Griffanti, L., & Smith, S. M. (2014). Automatic denoising of functional MRI data: Combining independent component analysis and hierarchical fusion of classifiers. NeuroImage, 90, 449–468. https://doi.org/10.1016/j.neuroimage.2013.11.046

Sato, J. R., Biazoli, C. E., Salum, G. A., Gadelha, A., Crossley, N., Vieira, G., Zugman, A., Picon, F. A., Pan, P. M., Hoexter, M. Q., Amaro, E., Anés, M., Moura, L. M., Del’Aquilla, M. A. G., McGuire, P., Rohde, L. A., Miguel, E. C., Jackowski, A. P., & Bressan, R. A. (2018). Association between abnormal brain functional connectivity in children and psychopathology: A study based on graph theory and machine learning. The World Journal of Biological Psychiatry, 19(2), 119–129. https://doi.org/10.1080/15622975.2016.1274050

Schaefer, A., Kong, R., Gordon, E. M., Laumann, T. O., Zuo, X.-N., Holmes, A. J., Eickhoff, S. B., & Yeo, B. T. T. (2018). Local-Global Parcellation of the Human Cerebral Cortex from Intrinsic Functional Connectivity MRI. Cerebral Cortex, 28(9), 3095–3114. https://doi.org/10.1093/cercor/bhx179

Sha, Z., Wager, T. D., Mechelli, A., & He, Y. (2019). Common Dysfunction of Large-Scale Neurocognitive Networks Across Psychiatric Disorders. Biological Psychiatry, 85(5), 379–388. https://doi.org/10.1016/j.biopsych.2018.11.011

Shirer, W. R., Jiang, H., Price, C. M., Ng, B., & Greicius, M. D. (2015). Optimization of rs-fMRI pre-processing for enhanced signal-noise separation, test-retest reliability, and group discrimination. NeuroImage, 117, 67–79.

Sydnor, V. J., Larsen, B., Bassett, D. S., Alexander-Bloch, A., Fair, D. A., Liston, C., Mackey, A. P., Milham, M. P., Pines, A., & Roalf, D. R. (2021). Neurodevelopment of the association cortices: Patterns, mechanisms, and implications for psychopathology. Neuron, 109(18), 2820–2846.

Uddin, M. N., Figley, T. D., Solar, K. G., Shatil, A. S., & Figley, C. R. (2019). Comparisons between multi-component myelin water fraction, T1w/T2w ratio, and diffusion tensor imaging measures in healthy human brain structures. Scientific reports, 9(1), 1-17.

van den Heuvel, M. P., & Sporns, O. (2019). A cross-disorder connectome landscape of brain dysconnectivity. Nature Reviews Neuroscience, 20(7), 435–446. https://doi.org/10.1038/s41583-019-0177-6

Vanes, L. D., & Dolan, R. J. (2021). Transdiagnostic neuroimaging markers of psychiatric risk: A narrative review. NeuroImage: Clinical, 30, 102634. https://doi.org/10.1016/j.nicl.2021.102634

Voldsbekk, I., Kjelkenes, R., Wolfers, T., Dahl, A., Lund, M. J., Kaufmann, T., Fernandez-Cabello, S., de Lange, A.-M. G., Tamnes, C. K., Andreassen, O. A., Westlye, L. T., & Alnæs, D. (2023). Shared pattern of impaired social communication and cognitive ability in the youth brain across diagnostic boundaries. Developmental Cognitive Neuroscience, 60, 101219. https://doi.org/10.1016/j.dcn.2023.101219

Woolrich, M. W., Ripley, B. D., Brady, M., & Smith, S. M. (2001). Temporal Autocorrelation in Univariate Linear Modeling of FMRI Data. NeuroImage, 14(6), 1370–1386. https://doi.org/10.1006/nimg.2001.0931

Xia, C. H., Ma, Z., Ciric, R., Gu, S., Betzel, R. F., Kaczkurkin, A. N., Calkins, M. E., Cook, P. A., García de la Garza, A., Vandekar, S. N., Cui, Z., Moore, T. M., Roalf, D. R., Ruparel, K., Wolf, D. H., Davatzikos, C., Gur, R. C., Gur, R. E., Shinohara, R. T., Bassett, D. S., & Satterthwaite, T. D. (2018). Linked dimensions of psychopathology and connectivity in functional brain networks. Nature Communications, 9(1), 3003. https://doi.org/10.1038/s41467-018-05317-y

Xia, M., Wang, J., & He, Y. (2013). BrainNet Viewer: A Network Visualization Tool for Human Brain Connectomics. PLoS ONE, 8(7), e68910. https://doi.org/10.1371/journal.pone.0068910

