## Supplementary material for "Delineating disorder-general and disorder-specific dimensions of psychopathology from functional brain networks in a developmental clinical sample"

**Content**


- Supplementary Figure 1. Sample flow chart.
- Supplementary Figure 2. Percentage missing data in each parcel
- Supplementary Figure 3. Correlations between 60% voxels and full 100% voxels in each parcel
- Supplementary Figure 4. Symptom-based PLS rerun without regressing out age and sex
- Supplementary Figure 5. Correlations between edges and symptom data by ethnic background
- Supplementary Figure 6. Correlations between edges and symptom data by median-split of household income
- Supplementary Figure 7. Correlations between edges and symptom data by IQ ± 70
- Supplementary Figure 8. Correlations between edges and symptom data by medication use
- Supplementary Figure 9. Scree plot of percent cross-block covariance explained in symptom-based PLS
- Supplementary Figure 10. Scatter plots of significant LVs in symptom-based PLS
- Supplementary Figure 11. Edges associated with symptom based PLS dimensions LV2-LV6
- Supplementary Figure 12. Edges specific to no diagnosis vs all diagnoses
- Supplementary Figure 13. Weights on each LV by diagnostic category and no. of diagnoses
- Supplementary Figure 14. Replication of symptom-based PLS between discovery and replication sample
- Supplementary Figure 15. Replication of diagnosis-based PLS between discovery and replication sample
- Supplementary Figure 16. Replication of diagnosis-specific PLS between discovery and replication sample
- Supplementary Table 1. Overview of contrasts used in the non-rotated diagnosis-specific PLS
- Supplementary Table 2. Effect of age and sex on symptom-based PLS
- Supplementary Table 3. Linear models of associations of each diagnosis with symptom and connectivity weights
- Supplementary Table 4. Linear models of associations of age, sex, and number of diagnoses (0-10) with symptom and connectivity weights on symptom-based PLS LV1
- Supplementary Table 5. Linear models of associations of age, sex, and number of diagnoses (1-10) with symptom and connectivity weights on symptom-based PLS LV1

**Supplementary Figure 1. Sample flow chart**

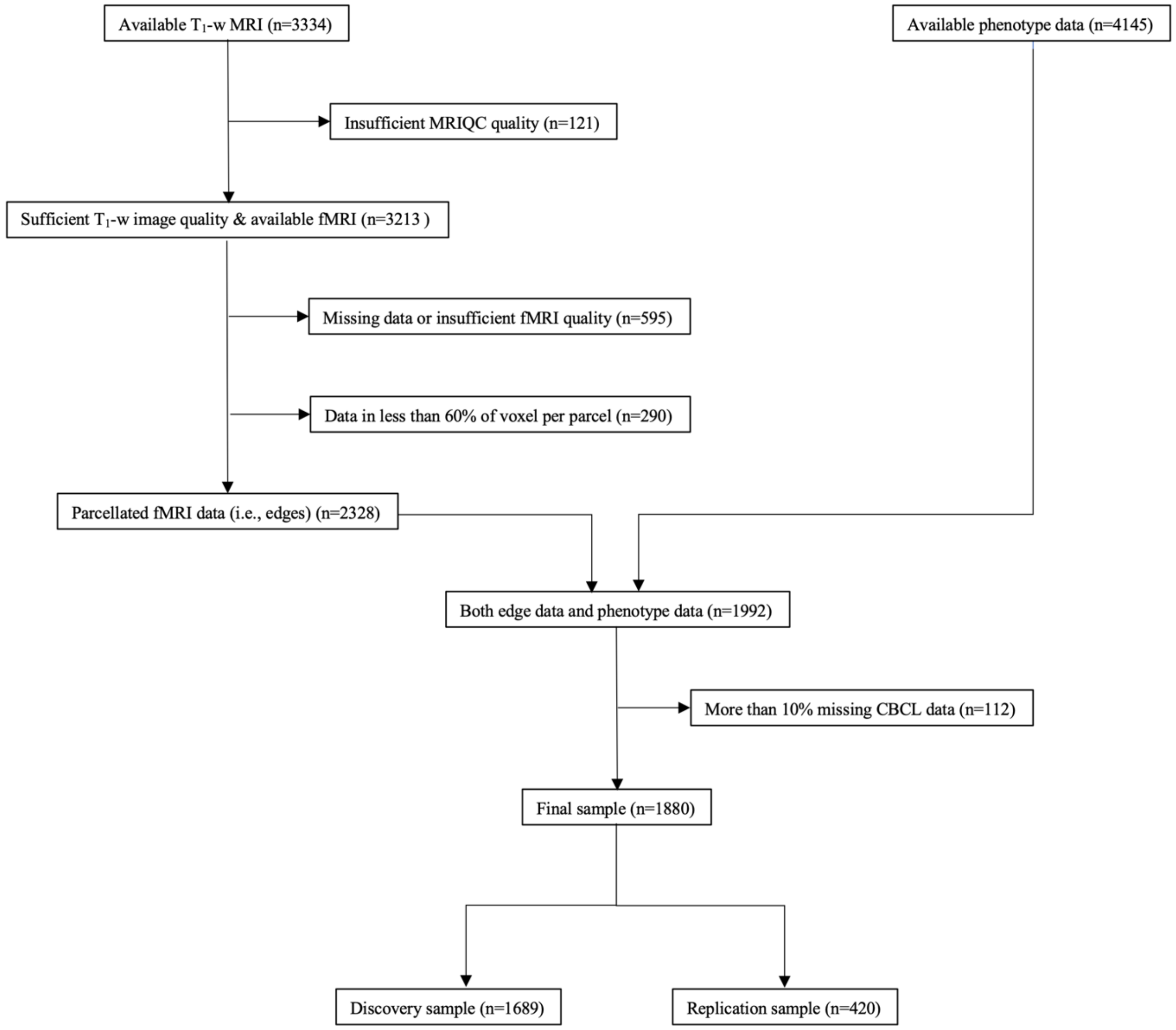

**Supplementary Figure 2. Percentage missing data in each parcel**

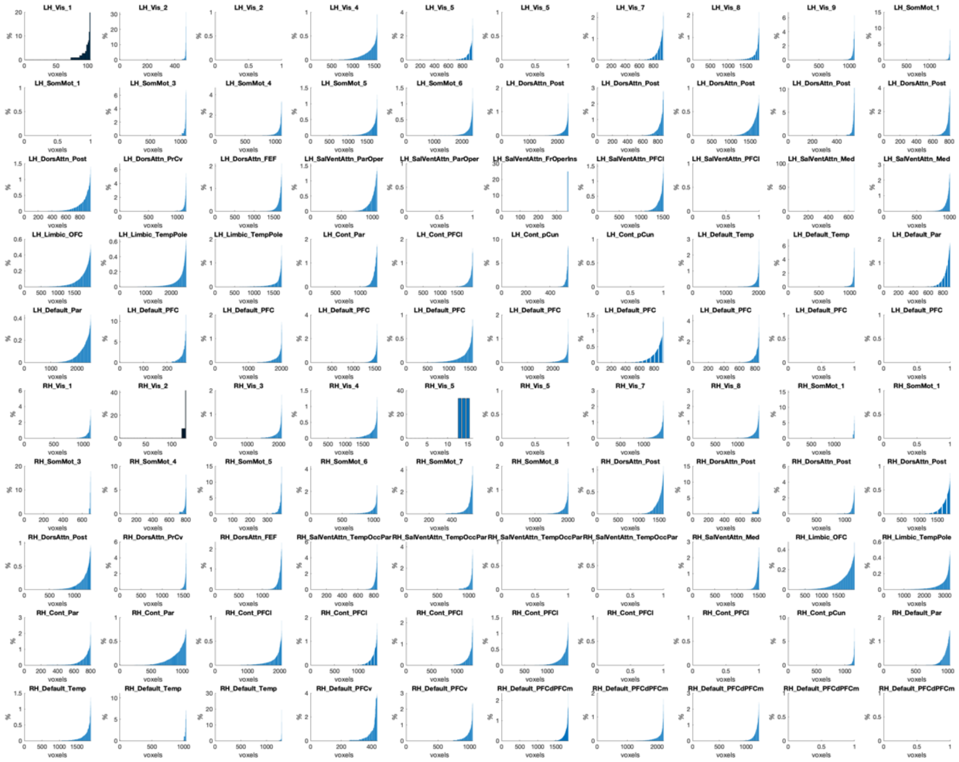

**Supplementary Figure 3. Correlations between 60% voxels and full 100% voxels in each parcel**

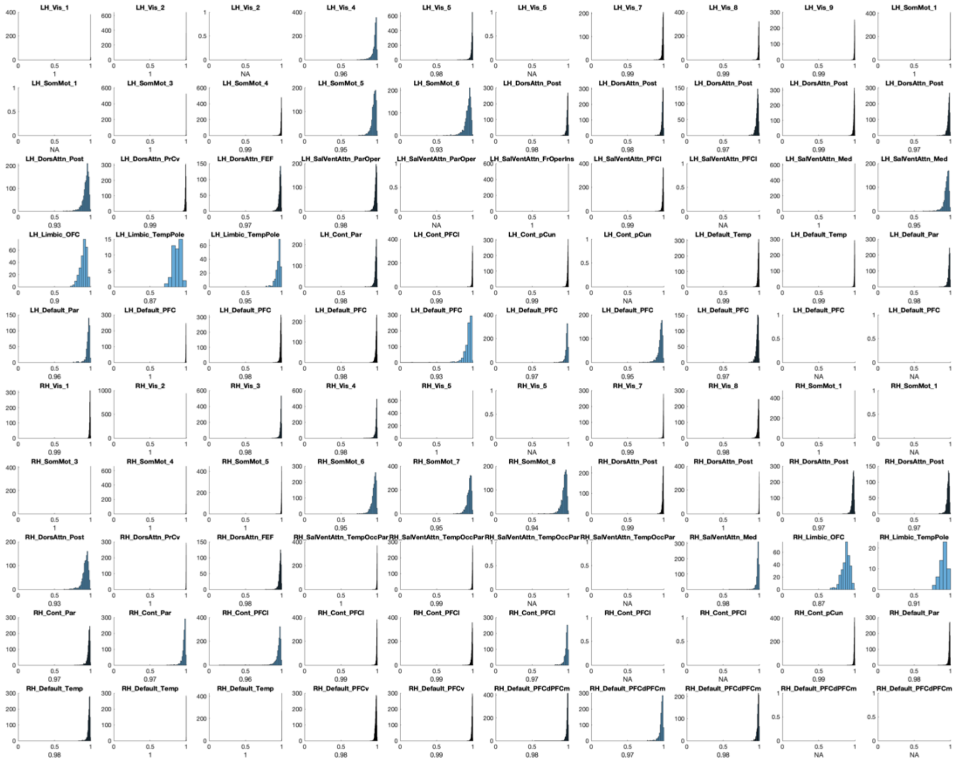

### Supplementary Figure 4. Symptom-based PLS rerun without regressing out age and sex

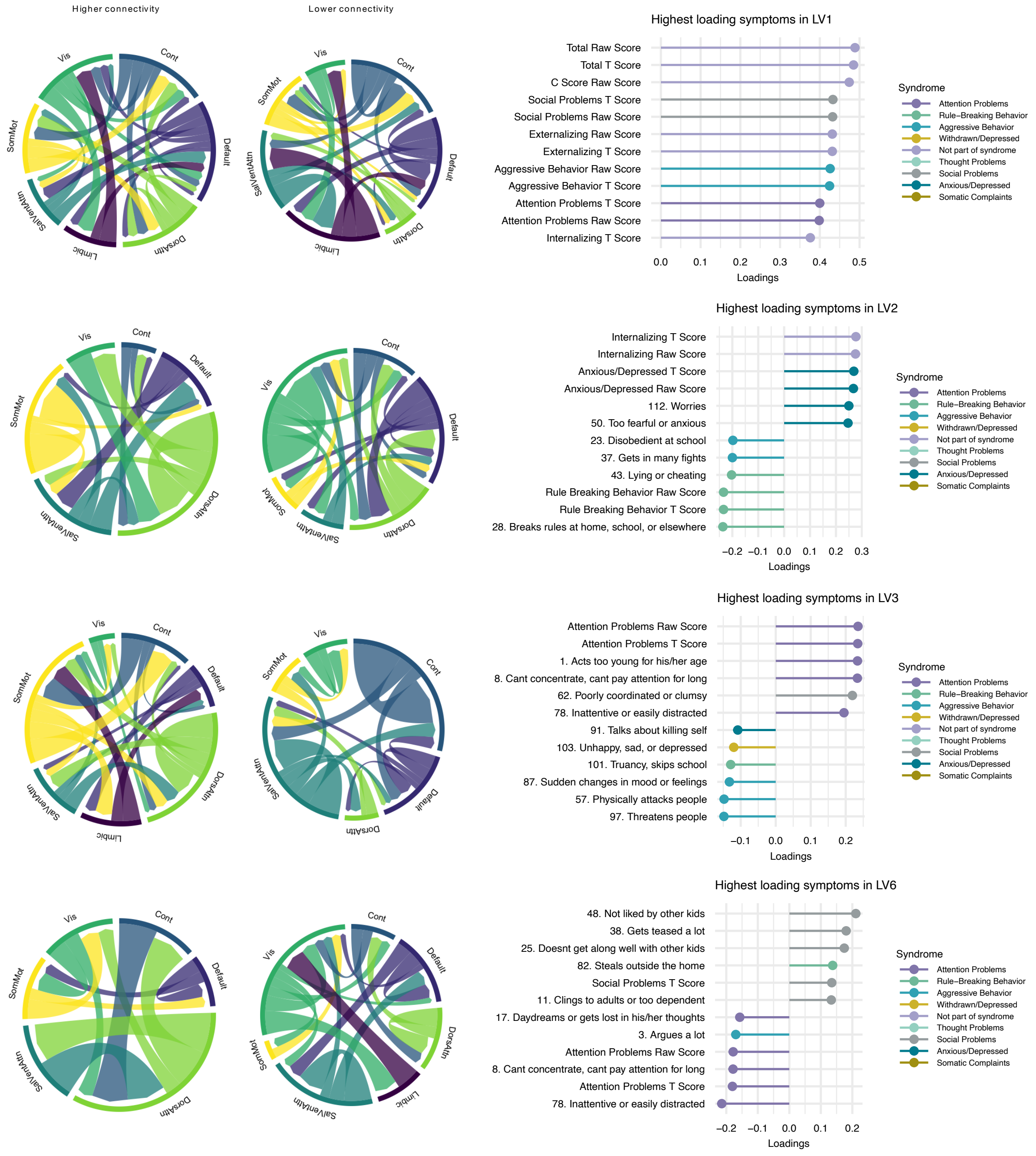

### Supplementary Figure 5. Correlations between edges and symptom data by ethnic background

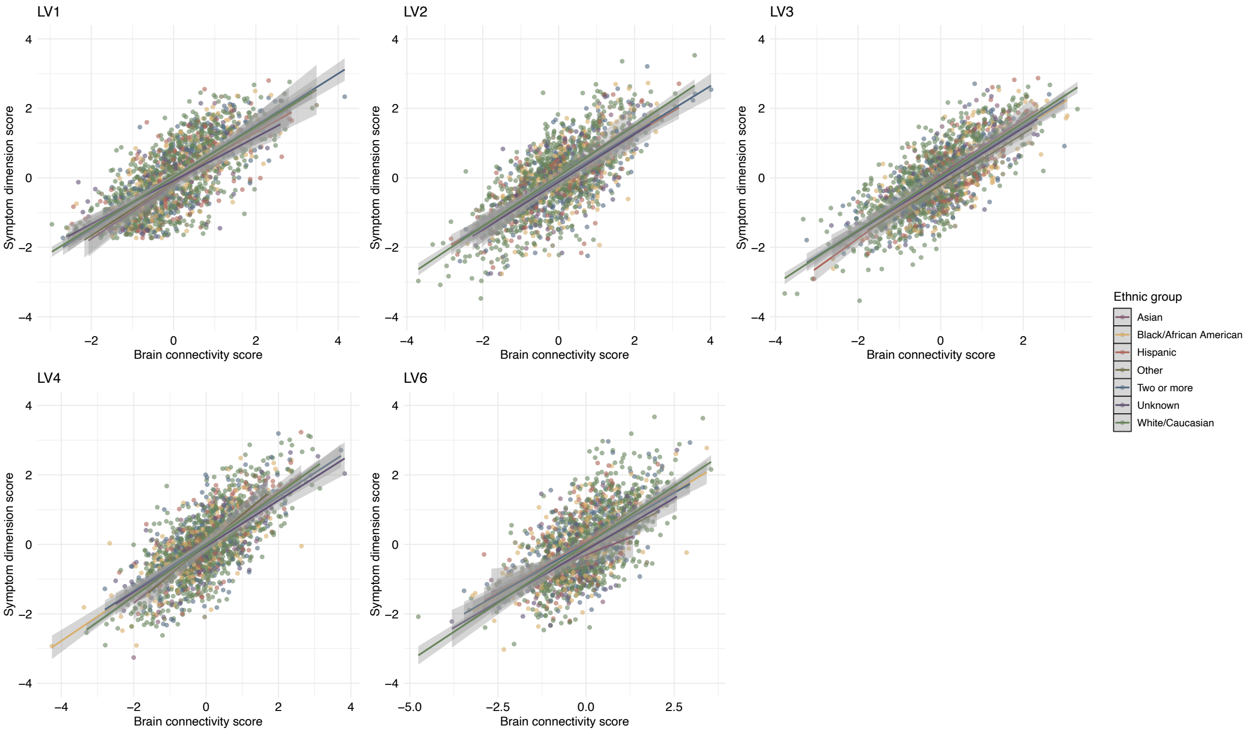

#### Supplementary Figure 6. Correlations between edges and symptom data by median-split of household income

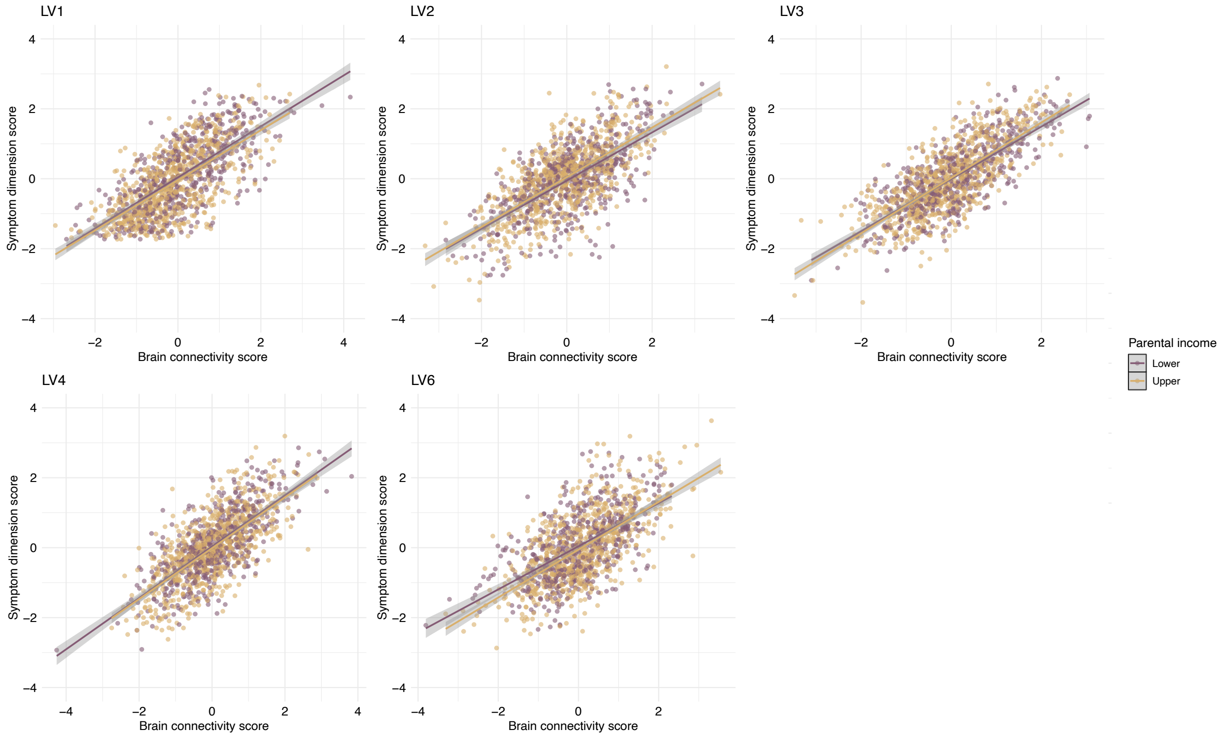

#### Supplementary Figure 7. Correlations between edges and symptom data by IQ ± 70

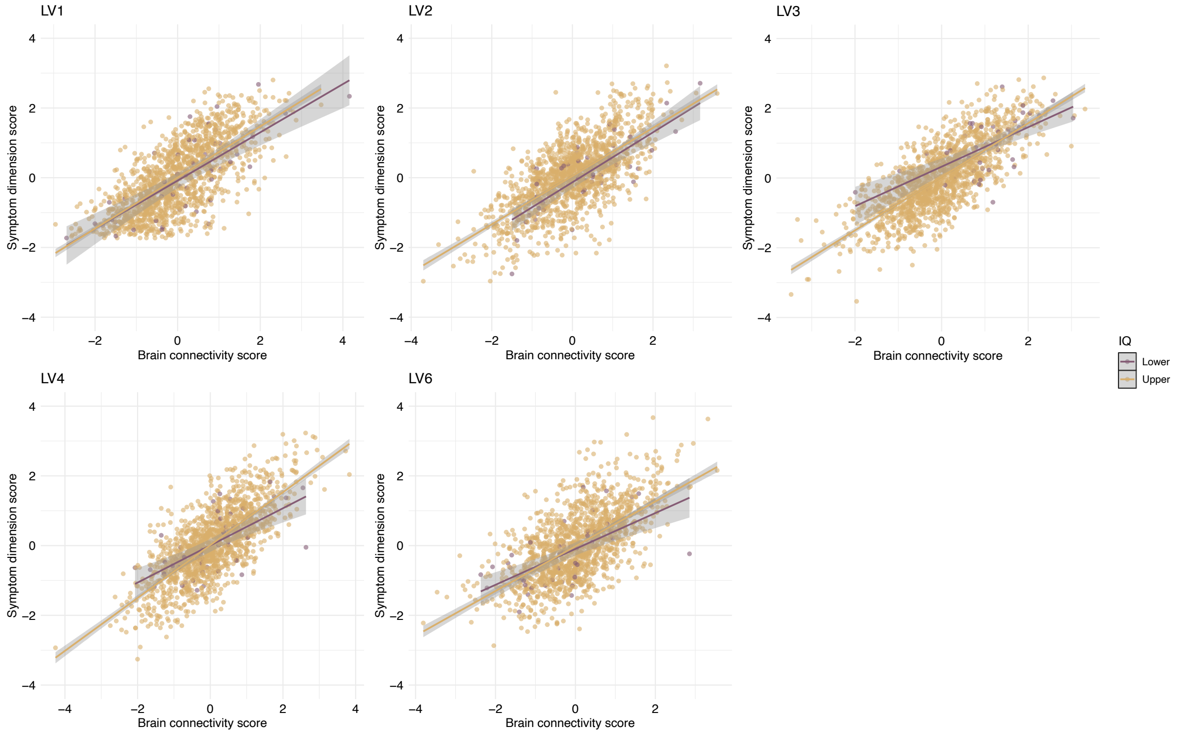

### Supplementary Figure 8. Correlations between edges and medication use

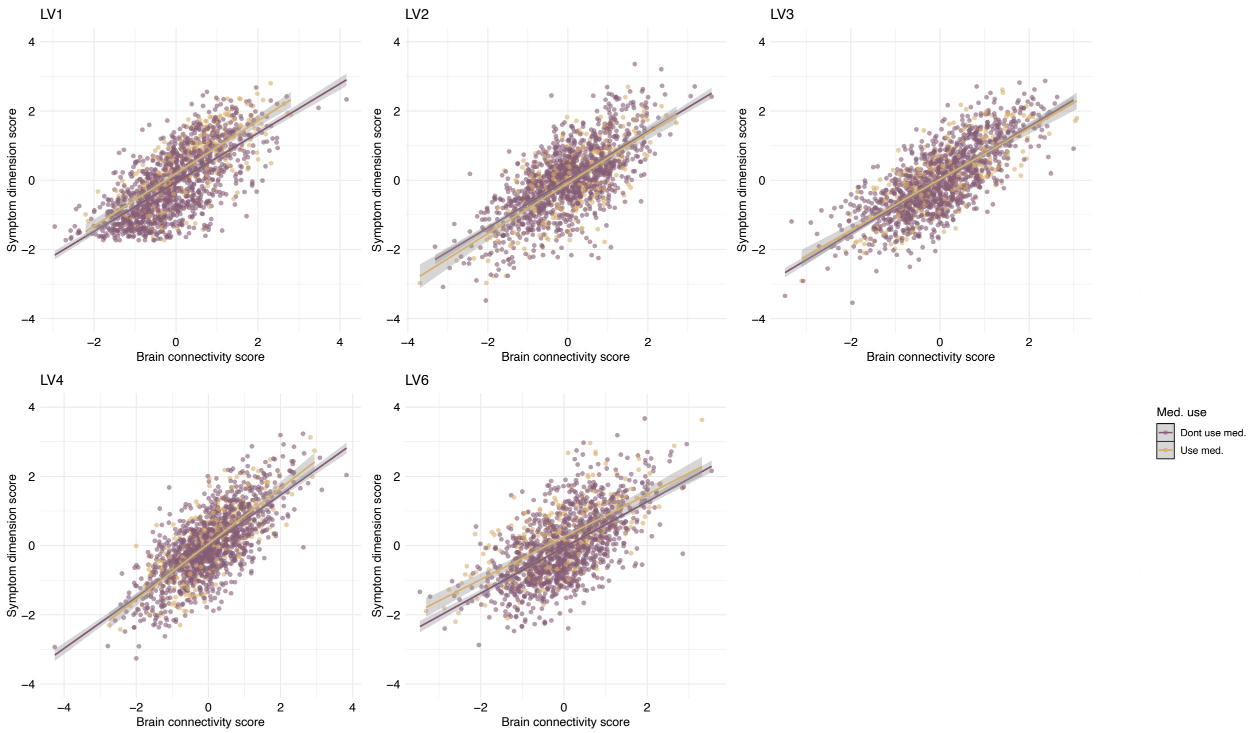

**Supplementary Figure 9. Scree plot of percent cross-block covariance explained in symptom-based PLS**

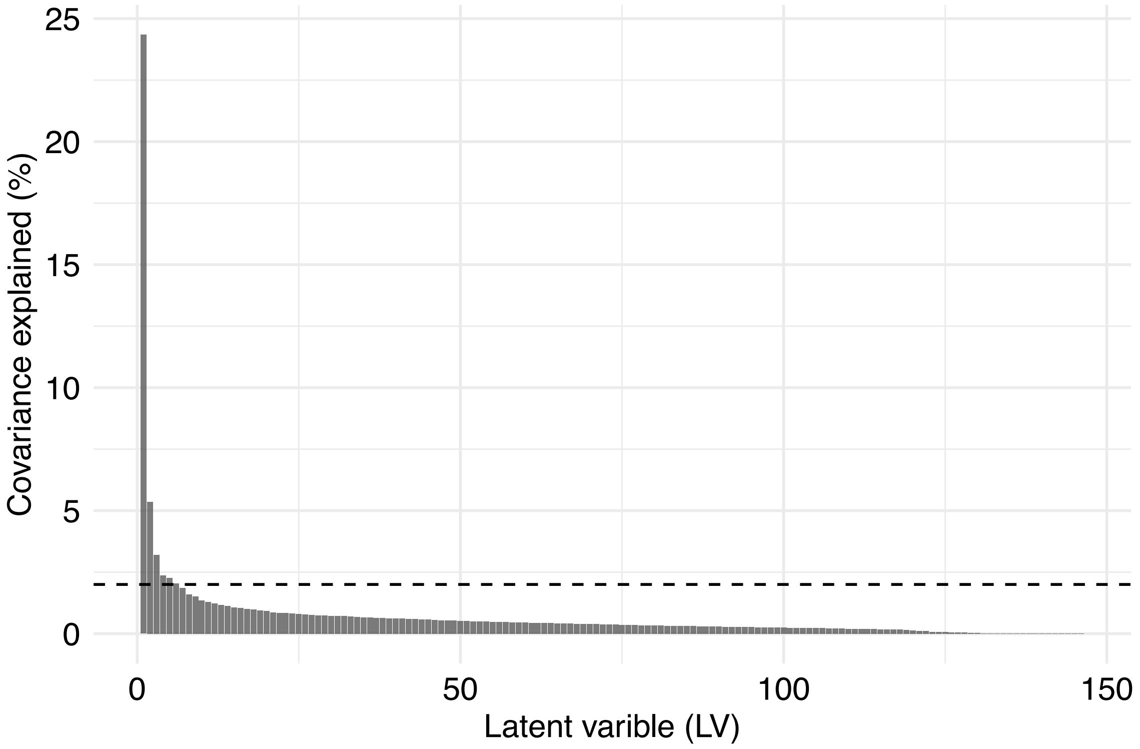

### Supplementary Figure 10. Scatter plots of significant LVs in symptom-based PLS

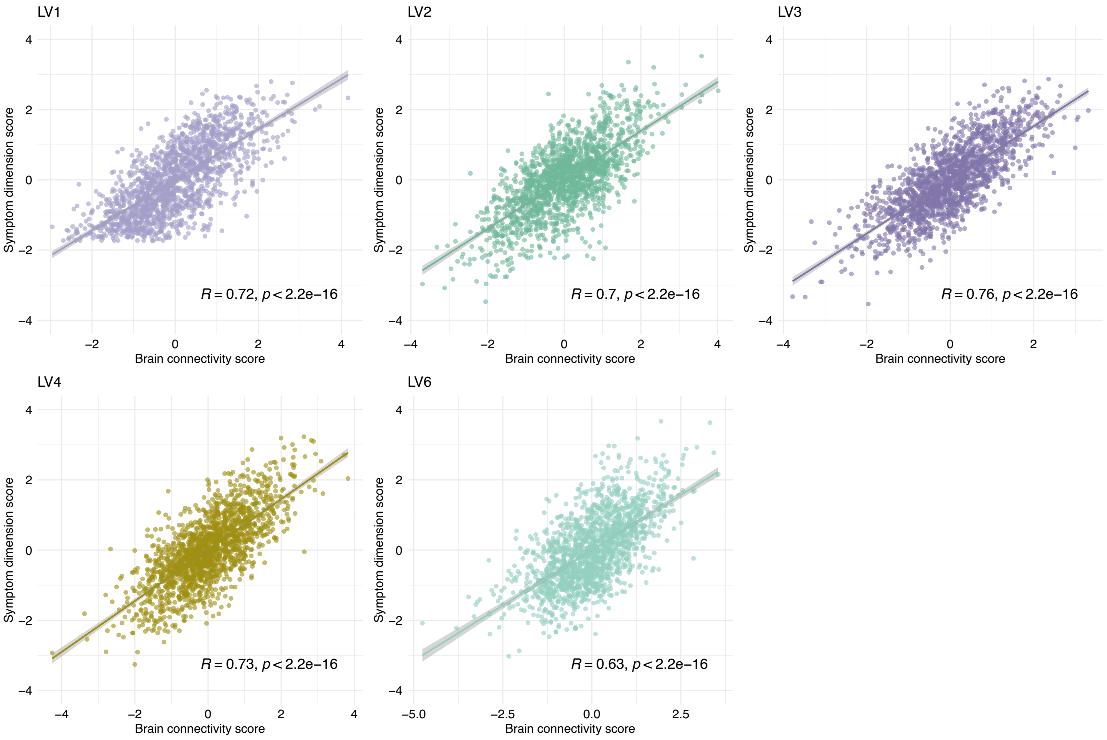

**Supplementary Figure 11. Edges associated with symptom based PLS dimensions LV2-LV6**

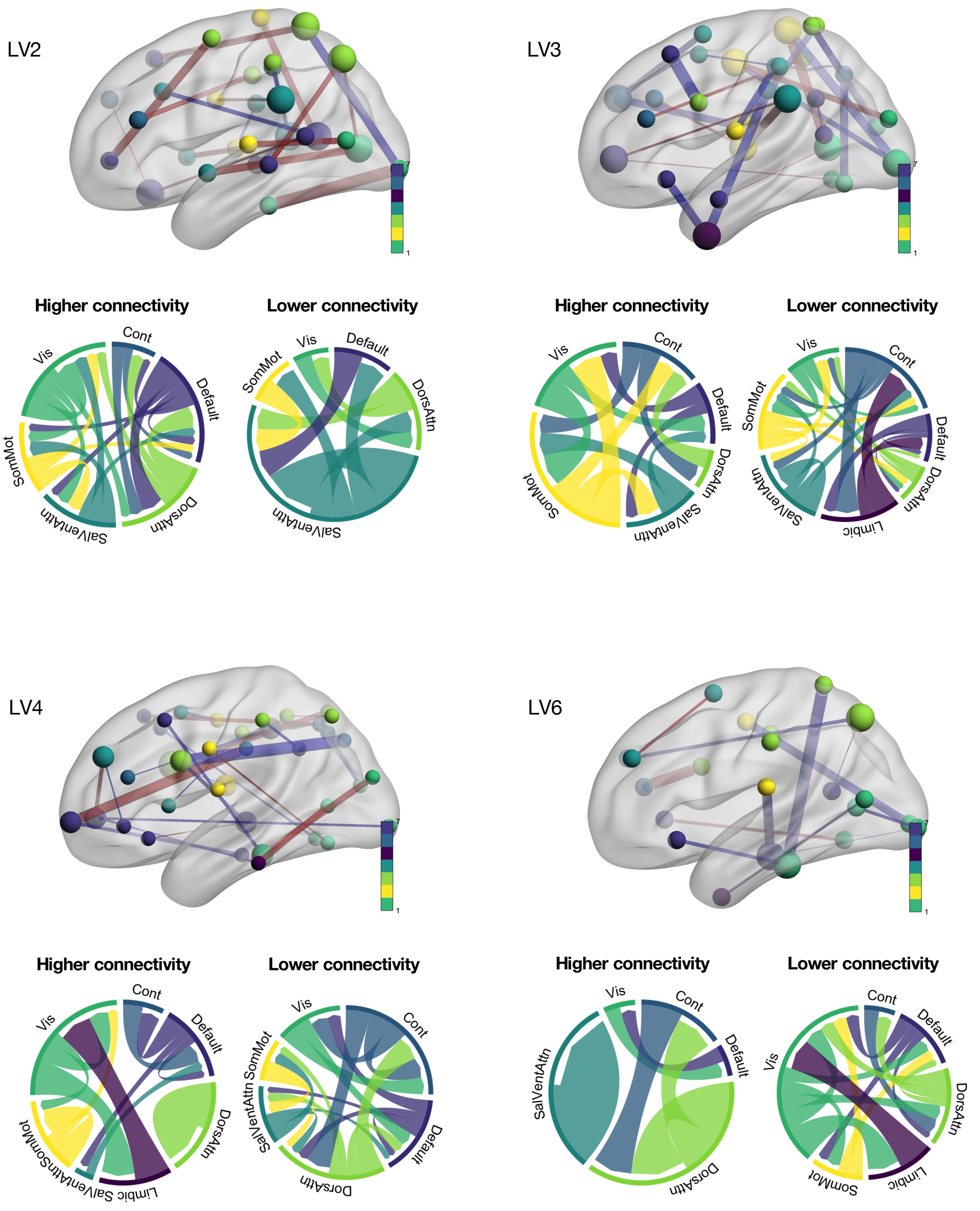

**Supplementary Figure 12. Edges specific to no diagnosis vs all diagnoses**

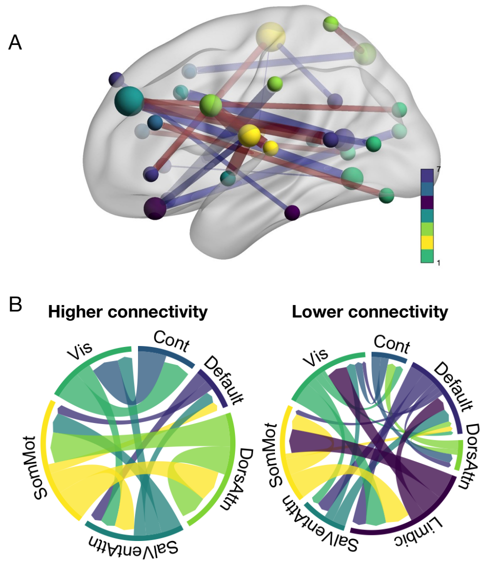

### Supplementary Figure 13. Weights on each LV by diagnostic category and no. of diagnoses

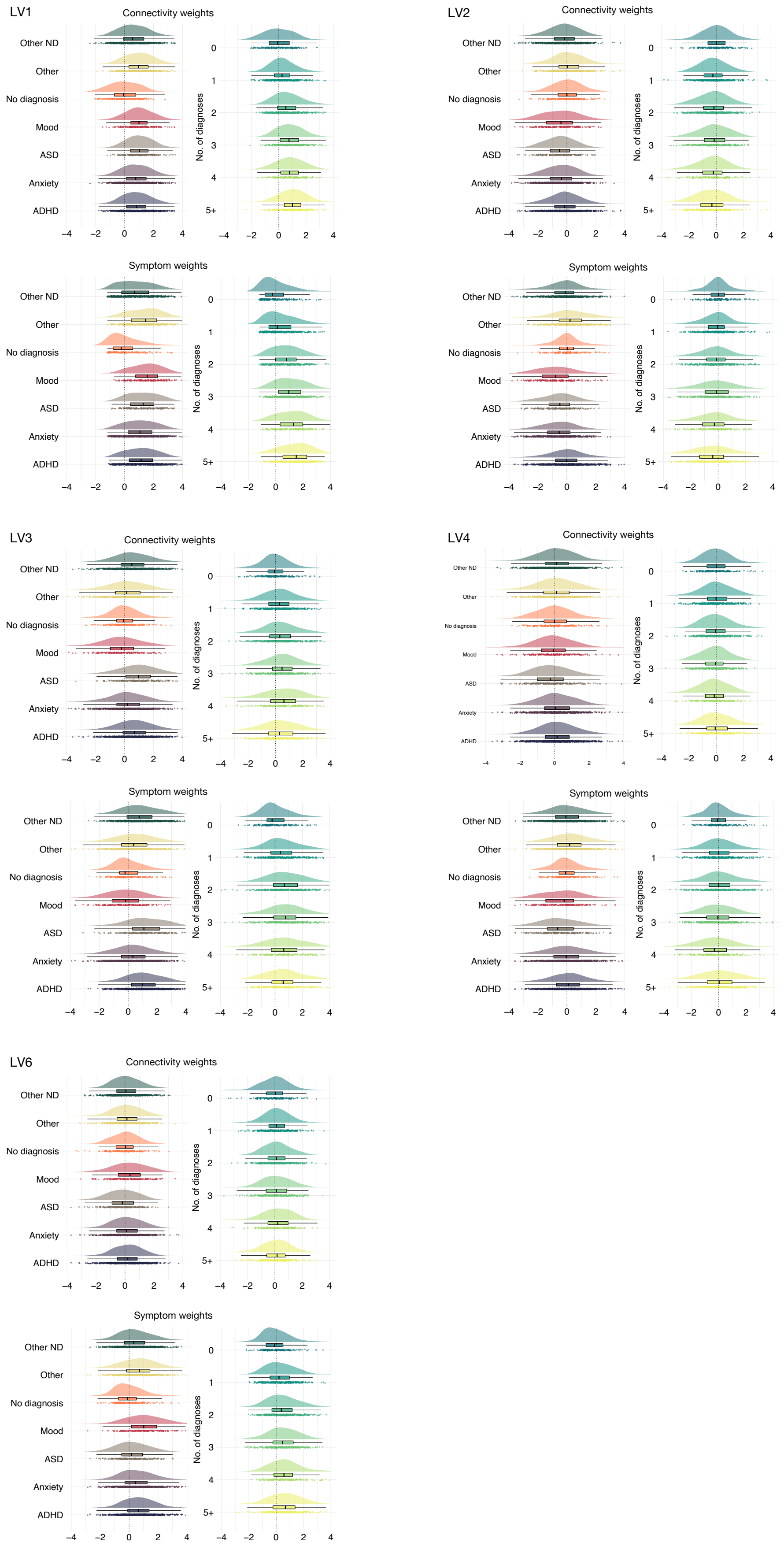

### Supplementary Figure 14. Replication of symptom-based PLS between discovery and replication sample

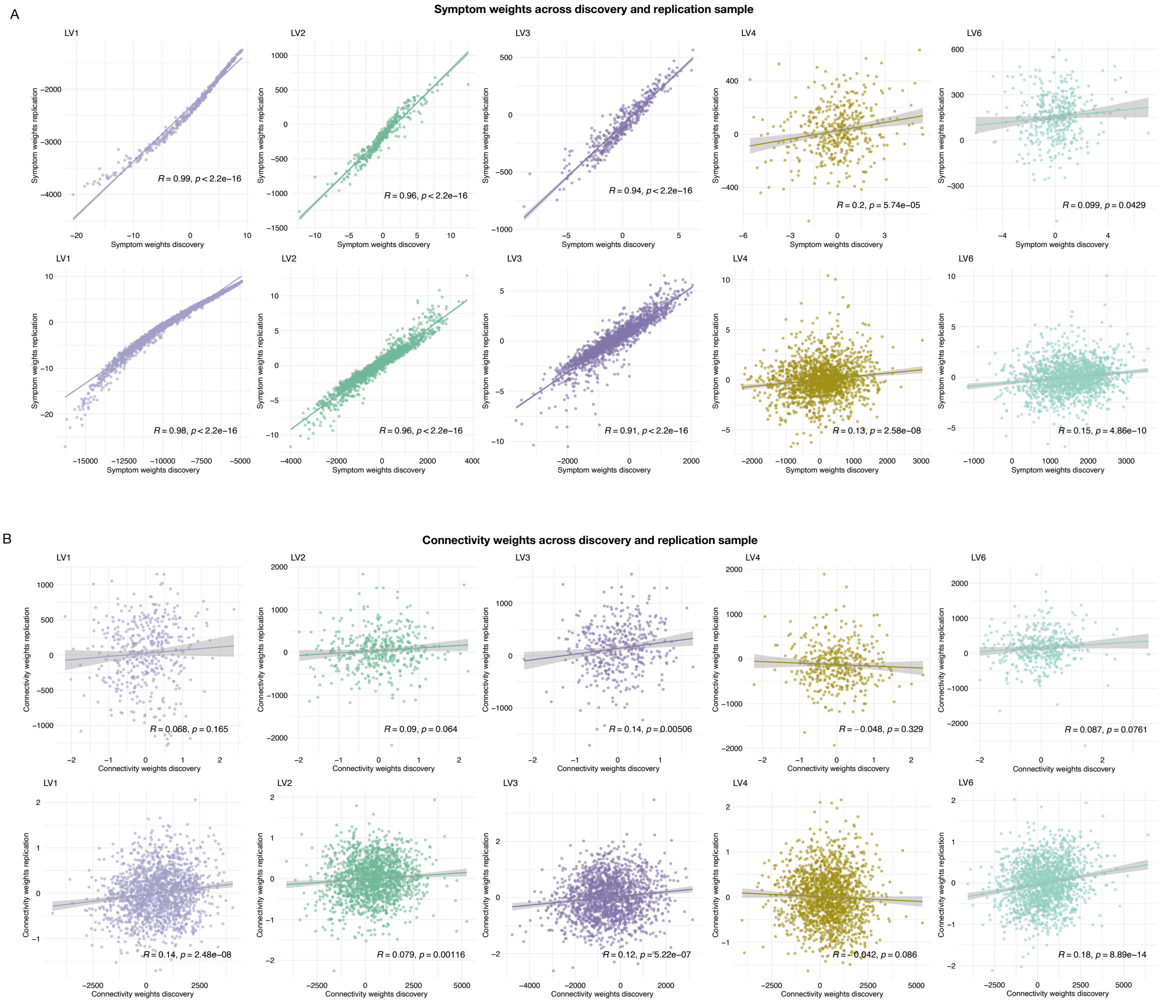

**Supplementary Figure 15. Replication of diagnosis-based PLS between discovery and replication sample**

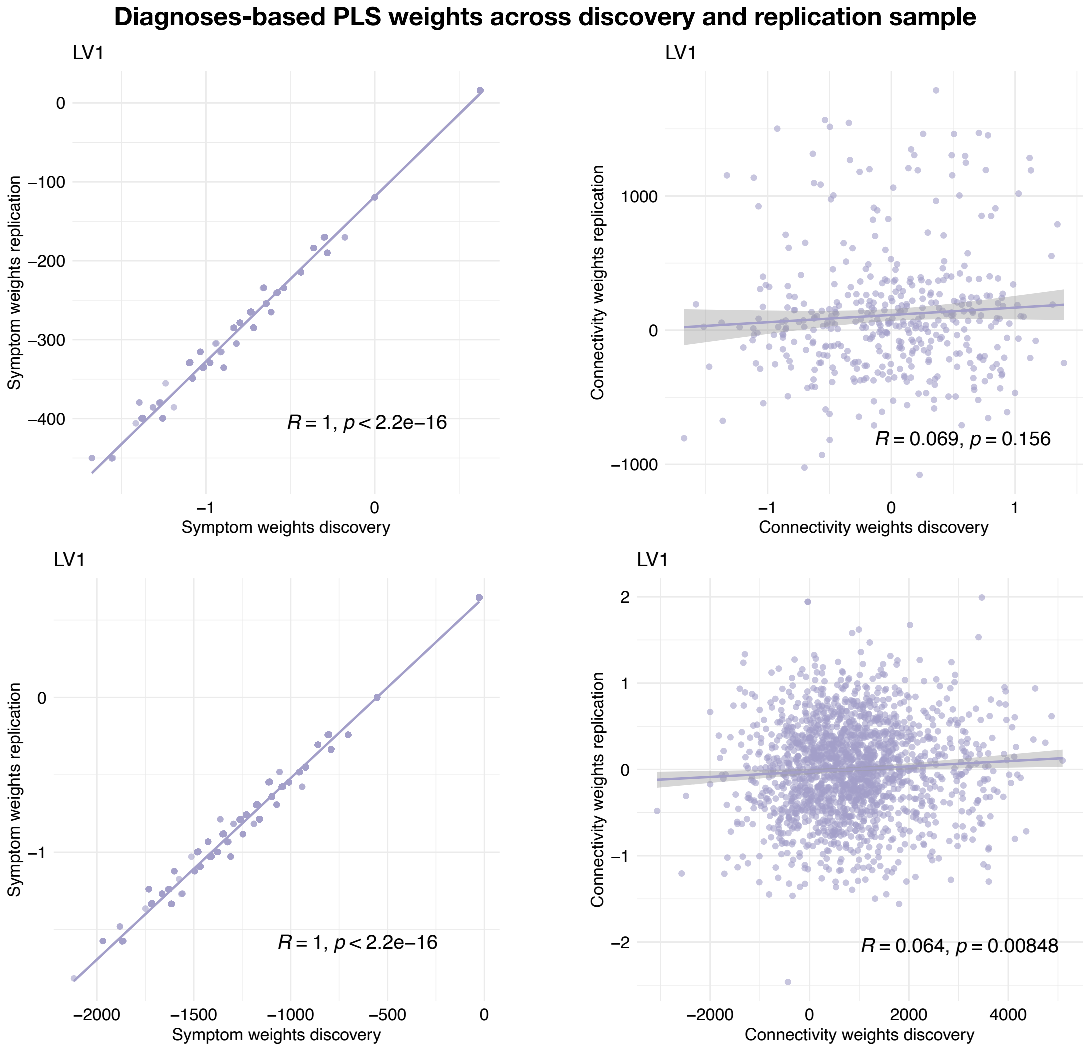

**Supplementary Figure 16. Replication of diagnosis-specific PLS between discovery and replication sample**

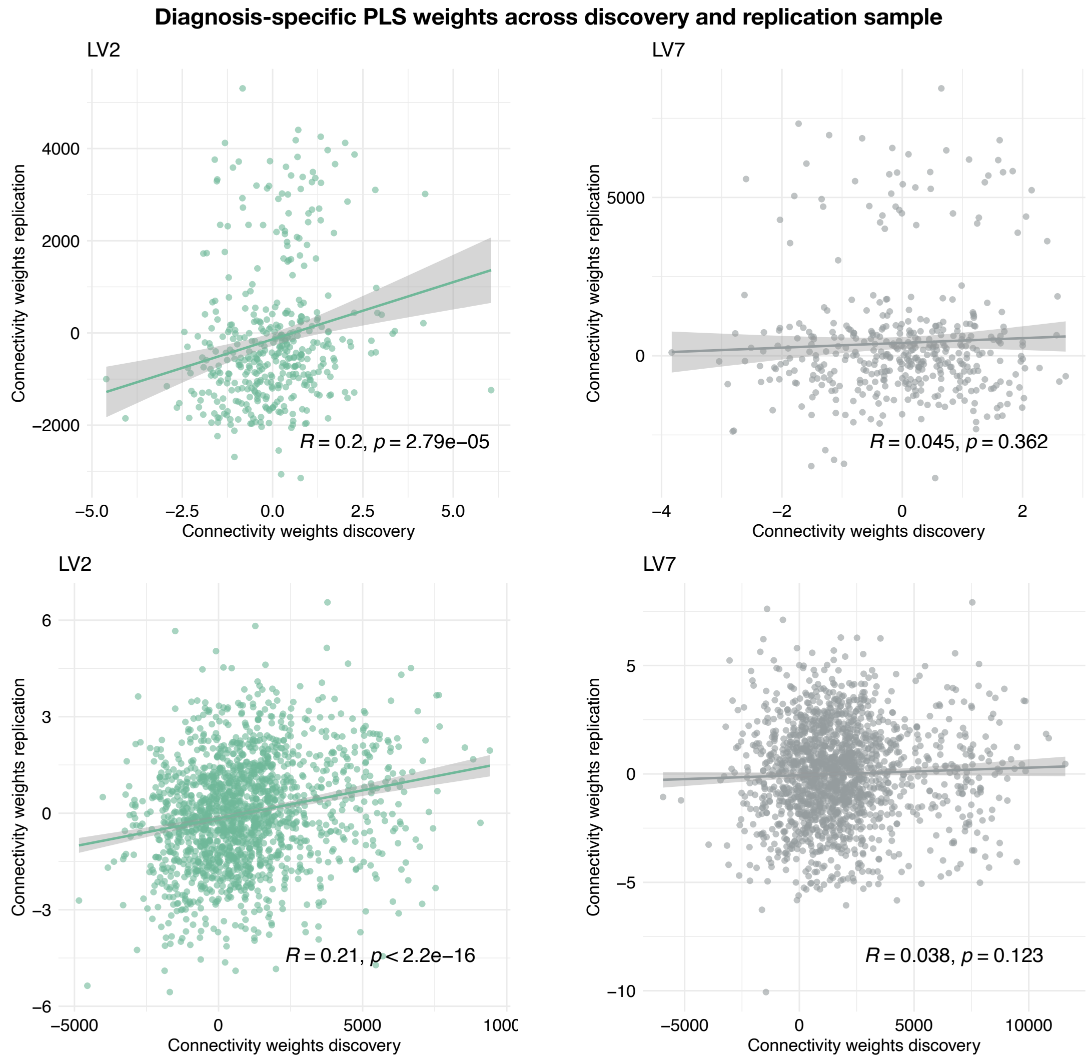

**Supplementary Table 1. Overview of contrasts used in the non-rotated diagnosis-specific PLS**

*Supplementary Table.* Overview of contrasts used in the non-rotated diagnosis-specific PLS.

| Contrasts | ADHD | ASD | Anxiety | Mood | Other ND | Other | No dx |
|---|---|---|---|---|---|---|---|
| 1 | 1 | -0.1667 | -0.1667 | -0.1667 | -0.1667 | -0.1667 | -0.1667 |
| 2 | -0.1667 | 1 | -0.1667 | -0.1667 | -0.1667 | -0.1667 | -0.1667 |
| 3 | -0.1667 | -0.1667 | 1 | -0.1667 | -0.1667 | -0.1667 | -0.1667 |
| 4 | -0.1667 | -0.1667 | -0.1667 | 1 | -0.1667 | -0.1667 | -0.1667 |
| 5 | -0.1667 | -0.1667 | -0.1667 | -0.1667 | 1 | -0.1667 | -0.1667 |
| 6 | -0.1667 | -0.1667 | -0.1667 | -0.1667 | -0.1667 | 1 | -0.1667 |
| 7 | -0.1667 | -0.1667 | -0.1667 | -0.1667 | -0.1667 | -0.1667 | 1 |

*Note.* PLS; partial least squares. ADHD; attention-deficit hyperactivity disorder. ASD; autism spectrum disorder. ND; neurodevelopmental. Dx; diagnosis.

**Supplementary Table 2. Effect of age and sex on symptom-based PLS**

*Supplementary Table.* Pearson correlations between PLS weights from the original, covariate-regressed symptom-based analysis and the repeated analysis without adjusting for age and sex. Both analyses are adjusted for scanner site, tSNR, and framewise displacement. Of note, LV4 was not itself significant in the PLS without adjusting for age and sex.

| | **Connectivity weights** | | **Symptom weights** | |
|---|---|---|---|---|
| | R | p | R | p |
| **LV1**: General p | 0.73 | 0 | 0.99 | 0 |
| **LV2**: Externalising-internalising | 0.84 | 0 | 0.99 | 0 |
| **LV3**: Neurodevelopment | 0.53 | 0 | 0.94 | 0 |
| **LV4**: Somatic complaints | 0.59 | 0 | 0.79 | 0 |
| **LV6**: Thought problems | 0.81 | 0 | 0.87 | 0 |

*Note.* PLS; partial least squares. tSNR; temporal signal-to-noise ratio. LV; latent variate.

**Supplementary Table 3. Linear models of associations of each diagnosis with symptom and connectivity weights**

*Supplementary Table.* Linear models of associations of each diagnosis with symptom and connectivity weights on symptom-based PLS LV1. Age, age$^2$, and sex are included as covariates

| | Symptom weights | | | | | | Connectivity weights | | | | | |
|---|---|---|---|---|---|---|---|---|---|---|---|---|
| | ADHD | ASD | Anx. | Mood | Other ND | Other | ADHD | ASD | Anx. | Mood | Other ND | Other |
| Age | .55 | 1.20 | .61 | -.74 | 2.19 | 2.72 | -.52 | .17 | -.99 | -2.37 | 1.36 | .94 |
| | (1.06) | (1.03) | (1.08) | (1.12) | (1.11) | (1.07) | (.97) | (.95) | (.98) | (1.06) | (.99) | (.98) |
| | p = .61 | p = .25 | p = .58 | p = .51 | p = .05 | p = .02 | p = .60 | p = .86 | p = .32 | p = .03 | p = .18 | p = .34 |
| Age$^2$ | .94 | -.33 | -.22 | .13 | 1.57 | -.12 | 2.47 | 1.49 | 2.35 | 2.69 | 2.91 | 2.33 |
| | (1.07) | (1.03) | (1.08) | (1.02) | (1.11) | (1.07) | (.98) | (.95) | (.98) | (.96) | (1.00) | (.98) |
| | p = .38 | p = .76 | p = .84 | p = .90 | p = .16 | p = .92 | p = .02 | p = .12 | p = .02 | p = .01 | p = .004 | p = .02 |
| Sex | -.04 | -.09 | .07 | .07 | .10 | .09 | -.08 | -.08 | -.03 | .01 | .02 | .07 |
| | (.07) | (.11) | (.07) | (.10) | (.07) | (.09) | (.06) | (.11) | (.07) | (.10) | (.07) | (.08) |
| | p = .54 | p = .45 | p = .35 | p = .50 | p = .17 | p = .30 | p = .21 | p = .45 | p = .70 | p = .93 | p = .79 | p = .37 |
| ADHD | 1.16 | | | | | | .86 | | | | | |
| | (.09) | | | | | | (.08) | | | | | |
| | p = 0.00 | | | | | | p = 0.00 | | | | | |
| ASD | | 1.29 | | | | | | 1.07 | | | | |
| | | (.11) | | | | | | (.10) | | | | |
| | | p = 0.00 | | | | | | p = 0.00 | | | | |
| Anxiety | | | 1.07 | | | | | | .82 | | | |
| | | | (.09) | | | | | | (.08) | | | |
| | | | p = 0.00 | | | | | | p = 0.00 | | | |
| Mood | | | | 1.53 | | | | | | 1.14 | | |
| | | | | (.11) | | | | | | (.11) | | |
| | | | | p = 0.00 | | | | | | p = 0.00 | | |
| Other ND | | | | | .76 | | | | | | .62 | |
| | | | | | (.09) | | | | | | (.08) | |
| | | | | | p = 0.00 | | | | | | p = 0.00 | |
| Other | | | | | | 1.35 | | | | | | .99 |
| | | | | | | (.09) | | | | | | (.09) |
| | | | | | | p = 0.00 | | | | | | p = 0.00 |
| Constant | .02 | .05 | -.03 | -.05 | -.07 | -.05 | .03 | .04 | .001 | -.07 | -.02 | -.05 |
| | (.08) | (.10) | (.09) | (.09) | (.09) | (.09) | (.08) | (.09) | (.08) | (.09) | (.08) | (.08) |

| | | | | | | | | | | | | |
|---|---|---|---|---|---|---|---|---|---|---|---|---|
| | p = .83 | p = .60 | p = .71 | p = .57 | p = .46 | p = .57 | p = .68 | p = .69 | p = .99 | p = .44 | p = .76 | p = .59 |
| Obs. | 1,105 | 405 | 895 | 381 | 958 | 610 | 1,105 | 405 | 895 | 381 | 958 | 610 |
| Adj. $R^2$ | .14 | .28 | .14 | .35 | .07 | .26 | .10 | .23 | .10 | .23 | .06 | .19 |
| Resid. Std. Error | 1.06 (df = 1100) | 1.03 (df = 400) | 1.07 (df = 890) | 1.01 (df = 376) | 1.11 (df = 953) | 1.07 (df = 605) | .97 (df = 1100) | .95 (df = 400) | .98 (df = 890) | .96 (df = 376) | .99 (df = 953) | .98 (df = 605) |

*Note.* Beta coefficient (standard error). ADHD; attention-deficit hyperactivity disorders. Anx; anxiety. ND; neurodevelopmental disorders.

**Supplementary Table 4. Linear models of associations of age, sex, and number of diagnoses (0-10) with symptom and connectivity weights on symptom-based PLS LV1**

*Supplementary Table.* Linear models of associations of age, sex, and number of diagnoses (0-10) with symptom and connectivity weights on symptom-based PLS LV1. Number of diagnoses included as a continuous variable.

| | **Symptom weights** | **Connectivity weights** |
|---|---|---|
| Age | -.66 (.95) | -1.36 (.97) |
| | p = .49 | p = .17 |
| $Age^2$ | .08 (.94) | 2.00 (.97) |
| | p = .94 | p = .04 |
| Sex | .06 (.05) | .002 (.05) |
| | p = .21 | p = .98 |
| No. of diagnoses (0-10) | .33 (.02) | .26 (.02) |
| | p = 0.00 | p = 0.00 |
| Constant | -.04 (.04) | -.001 (.04) |
| | p = .33 | p = .99 |
| Observations | 1,689 | 1,689 |
| Adjusted $R^2$ | .11 | .07 |
| Residual Std. Error (df = 1684) | .94 | .97 |

*Note.* Beta coefficient (standard error).

**Supplementary Table 5. Linear models of associations of age, sex, and number of diagnoses (1-10) with symptom and connectivity weights on symptom-based PLS LV1**

*Supplementary Table.* Linear models of associations of age, sex, and number of diagnoses (1-10) with symptom and connectivity weights on symptom-based PLS LV1. Number of diagnoses included as a continuous variable, excluding no diagnosis (0).

| | **Symptom weights** | **Connectivity weights** |
|---|---|---|
| Age | -1.21 (.96) | -1.37 (.97) |
| | p = .21 | p = .16 |
| $Age^2$ | -.24 (.95) | 1.12 (.96) |
| | p = .80 | p = .25 |
| Sex | .05 (.05) | -.01 (.05) |
| | p = .29 | p = .87 |
| No. of diagnoses (1-10) | .29 (.03) | .21 (.03) |
| | p = 0.00 | p = 0.00 |
| Constant | .01 (.04) | .05 (.04) |
| | p = .89 | p = .25 |
| Observations | 1,496 | 1,496 |
| Adjusted $R^2$ | .07 | .04 |
| Residual Std. Error (df = 1491) | .95 | .96 |

*Note.* Beta coefficient (standard error).
